# Metabolomic signature of weight loss and association with heart failure

**DOI:** 10.1101/2025.11.14.25340254

**Authors:** Nicholas Sunderland, Madeleine L. Smith, Alex McConnachie, Paul Welsh, Roy Taylor, Michael E. J. Lean, Chris A. Rogers, Jane M. Blazeby, Naveed Sattar, Lavinia Paternoster, R. Thomas Lumbers, Nicholas J. Timpson, Laura J. Corbin

## Abstract

**Background:** Obesity is a major risk factor for heart failure (HF), but the molecular mediators linking adiposity to HF remain unclear. The molecular mechanisms by which weight loss reduces the risk of HF are also unknown. Understanding these mechanisms could highlight potential therapeutic targets for all HF patients, including those who are normal weight. We aimed to identify a common metabolic perturbation profile by comparing different weight-loss interventions and to estimate their associations with HF using Mendelian randomisation (MR).

**Methods:** We first integrated mass spectrometry and nuclear magnetic resonance metabolomic profiling from two weight-loss interventions - a structured diet programme (DiRECT trial) and bariatric surgery (By-Band-Sleeve trial) - with estimates of the effect of life-time body mass index (BMI) exposure on metabolite levels through MR analyses, to identify a consistent BMI-metabolite signature across differing sources of BMI variation. We then assessed the impact of these BMI-metabolites on incident HF within a two sample MR framework.

**Results:** 1706 metabolites were analysed across three different sources of BMI variation: bariatric surgery, dietary intervention and life-time BMI exposure. 153 (9%) showed strong evidence for association with all three exposures with concordant direction of effect, predominantly comprising lipid fractions, lipoproteins, and amino acid metabolites. Among these metabolites, 44 (29%) had evidence of causal association with at least one HF subtype in MR. Notably, circulating levels of the non-lipid metabolites N-acetylglycine and asparagine were each inversely associated with BMI and with the risk of HF and HF with preserved ejection fraction risk, respectively. Both metabolites have previously been implicated in myocardial function and HF.

**Conclusions:** Our findings suggest that despite differences in the modality of weight loss delivery, there exists a consistent metabolomic profile coincident with weight change. Investigating the association of the identified metabolites with HF provides insights into molecular mediators of the effects of adiposity on HF and potential novel targets for therapeutic intervention.

## Introduction

Body mass index (BMI) is an anthropometric construct, often used as a proxy measure for adiposity and irrefutably associated with health.^1^ Of a swathe of downstream health outcomes associated with BMI, higher levels are recognised as a major risk factor for the development of heart failure (HF), more strongly to HF with preserved ejection fraction (HFpEF).^2^ Unlike the consistent declines seen in the rate of atherosclerotic cardiovascular (CV) disease in high-income countries through focus on traditional risk factors, such as blood pressure, hypercholesterolaemia and diabetes, HF incidence is now rising,^3^ with the rise in obesity thought to be partially responsible. Moreover, whilst the association between obesity and atherosclerotic CV disease is more modest and can be largely explained by these traditional CV mediators, the same is not true of HF and the exact routes from variation in BMI to clinical outcomes remains unclear.^3–5^

As a measure, BMI is a practical solution to the challenge of estimating excess adiposity at the population level, being derived from simple metrics and with simple associated guidelines.^6^ BMI is also the most accurate anthropometric measure. This underpins why BMI remains the most frequently used form of body composition assessment,^7^ however conceals the challenge of better understanding the link between adiposity and health outcomes. As an example, BMI does not differentiate between fat and lean mass, it is unable to precisely capture fat distribution and makes assumptions as to the multitude of combinations of physical presentation able to deliver equal numeric values in units of kg/m^2^.^8^ Alternative indices have been extensively investigated and measures such as waist hip ratio, or imaging derived adiposity parameters, can offer better predictive ability in certain scenarios, but ultimately are highly correlated with BMI and less scalable.^7,9,10^ In this context, therefore, whilst there is clear value in the application of simple and generic measures, there is a role for using intermediate biological traits to characterise the heterogeneity and nuance lying behind important axes of health risk. Theoretically, the outward phenotype may be less important than understanding the underlying biology that BMI summarises, not least because BMI is linked to such a diverse range of diseases, suggesting that distinct components of adiposity may be driving each of these associations.^11^

Efficient and cost effective metabolomic profiling studies offer the opportunity to further examine the underlying biology behind variation in BMI and responses to it. This can be achieved through precise molecular measurement of circulating small molecules which are themselves a by-product of cellular activity. Differences in circulating metabolite profiles between obese and normal weight individuals may highlight adiposity-related metabolic pathways; however, they may also capture coincident signals related to interventions, medication use, or bystander phenomena such as prevailing health. One strategy to distinguish confounding or coincident metabolic signals from adiposity-specific effects is to examine consistent signatures across contrasting study designs, each with independent (exogenous) sources of BMI variation.

Further to the characterisation of biological events involved in, or coincident with weight change, the molecular characterisation of adiposity can also be used to open up the examination of disease liability. This has been done by using prospective studies of outcomes such as cancer to dissect molecular intermediaries between BMI and disease.^12^ Additionally, natural genetic variation in these intermediaries allows for the dissection of potential causal pathways through Mendelian randomization (MR) analyses.

The aim of this study is to characterise the metabolic footprint of adiposity, leveraging separate sources of BMI variation to highlight consistent biology and consider implications for HF and its treatment. We hypothesise that the overlap between metabolite profiles of weight loss and that of HF (either through direct metabolomic profiling of HF patients or through instrumental variable MR analyses) can shed light on the components of adverse adiposity most important in the development of HF and therefore generate potentially novel targets for prevention and treatment of the condition.

## Methods

An overview of the study is presented in **Figure 1**.

**Figure 1.**
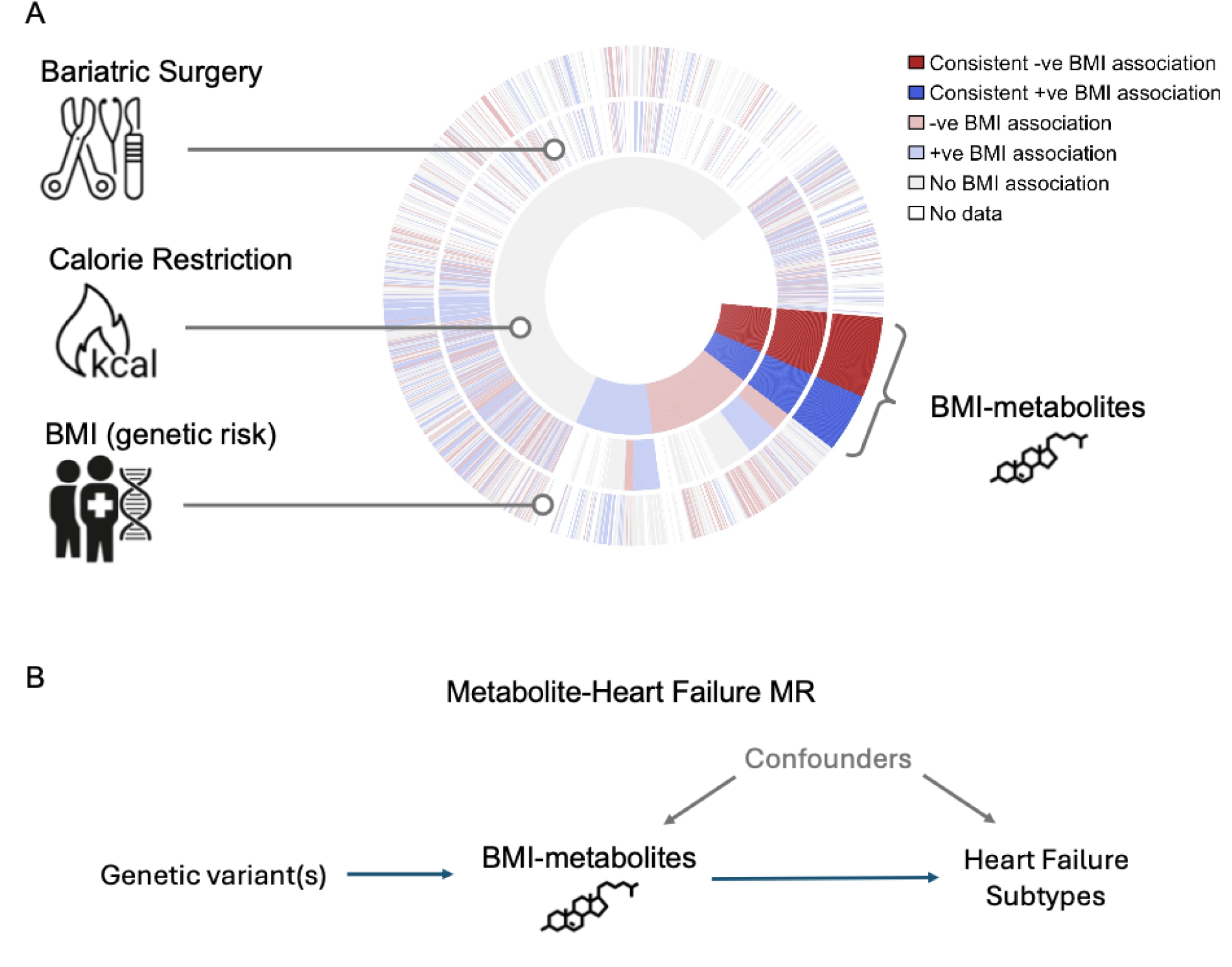
Analysis overview. (A) Circos plot showing metabolite associations with BMI across three independent sources of BMI variation: the inner ring represents dietary intervention, the middle ring bariatric surgery, and the outer ring population-level BMI variation assessed using Mendelian randomization (MR). Colours denote the direction of association (red, inverse; blue, positive; grey, missing data; bold colours represent consistent effects seen across all three sources). (B) Simplified directed acyclic graph (DAG) illustrating the MR framework used to test causal effects of circulating metabolite levels on heart failure risk.

### Study populations

To identify metabolites associated with adiposity, we examined the effects of two distinct weight-loss interventions - dietary intervention (Diabetes Remission Clinical Trial - DiRECT) and bariatric surgery (By-Band-Sleeve Trial) - on the circulating metabolome. The trials have been described in detail previously.^13–17^

Briefly, the DiRECT trial was a 2 year open-label, cluster randomised controlled trial (RCT) conducted at 49 primary care practices in Scotland and the Tyneside region of England between 25 July 2014 and 5 August 2016 (isrctn.org registration no. ISRCTN03267836). Individuals with type 2 diabetes mellitus (T2DM), a BMI of 27–45 kg/m^2^ and who were not receiving insulin, were randomised to either a weight management programme (intervention) or best-practice care by guidelines (control). The intervention comprised withdrawal of glucose-lowering and antihypertensive drugs, total diet replacement (3452–3569 kJ [825–853 kcal]/day formula diet for 3–5 months), stepped food reintroduction (2–8 weeks) and structured support for long-term weight-loss maintenance. Blood samples were available for metabolomic profiling at baseline and the endpoint was metabolic profiling at 12 months post-randomization. The intervention effect on metabolite levels was derived from the change seen in the intervention group, relative to the control group, over time. Ethics approval for DiRECT was granted by West 3 Ethics Committee in January 2014, with approvals by the National Health Service (NHS) health board areas in Scotland and clinical commissioning groups in Tyneside. All participants provided written informed consent.

The By-Band-Sleeve trial was a multi-site RCT designed to assess the most effective type of bariatric surgery (Roux-en-Y gastric bypass, laparoscopic adjustable gastric band, or sleeve gastrectomy) for weight loss and quality of life (EQ-5D-5L) as well as cost-effectiveness (isrctn.org registration no. ISRCTN00786323). Individuals with a BMI of ≥40 kg/m^2^, or ≥35 kg/m^2^ with other comorbidities such as T2DM, who were fit for general anaesthesia and surgery were recruited and randomised to one of the three types of bariatric surgery. Only individuals who eventually underwent surgery were included. Two sample collection efforts were made within By-Band-Sleeve – firstly, from participants in the main RCT and secondly from participants in a nested non-randomised (observational) sub-study. In both instances, samples were collected from patients at two time points. Ten of the twelve sites that participated in the main RCT collected samples for future research with blood samples available for metabolomic profiling at baseline and 36-months post-randomization. Seven of the twelve sites collected samples for future research in the non-randomised sub-study with blood samples available for metabolomic profiling at baseline and 12-months post-surgery. For the analysis presented here, this study is considered a single-arm (surgery) pre-post intervention study with the effect on metabolite levels derived from within-subject change over time. Ethical approval for BBS was granted by the Southwest Frenchay Research Ethics Committee (reference 11/SW/0248) and written informed consent was obtained from all participants. All samples were used, stored and disposed of in accordance with the Human Tissue Act 2004.

### Sample collection and metabolite data acquisition

Sample handling procedures have been described previously.^14,18^ Participants were asked to fast overnight before blood draw. All analysts were blinded to intervention status. Samples were sent for metabolomic profiling on the untargeted mass spectrometry (MS) Metabolon Global DIscovery Panel platform (Metabolon, Durham, NC, USA) and the ^1^H-NMR (NMR) Nightingale platform (Nightingale Health, Finland).

### Metabolite data preparation

Data relating to participants who withdrew consent during the study were removed from the analysis. Data quality was assessed locally using the *metaboprep* R package and predefined quality control (QC) metrics, as previously described.^14,19^ Samples with greater than 20% missingness, outlier samples with standard deviation (SD) >5 from the mean based on principal components, and samples > 5 SD from the total peak area (sum of metabolite levels for each individual at metabolites with no missingness), were identified and removed. Metabolites with >20% missingness were also removed.

QC procedures for the By-Band-Sleeve MS dataset lead to a total of 1410 samples with 952 metabolites for analysis, and for the NMR dataset 1386 samples with 250 metabolites. The same QC procedures in the DiRECT MS dataset lead to a total of 559 samples with 1276 metabolites, and for the NMR dataset 555 samples with 225 metabolites (**Supplemental Figure 1 & 2**).

MS datasets were transformed using a rank-based inverse normal transformation (INRT, where tied ranks were split by assigning random order). NMR datasets were transformed using Z-score normalisation, standardising each metabolite by subtracting the mean and dividing by the SD. Z-score normalisation was used as meaningful units can be preserved with NMR data whereas the MS data is semi-quantitative.

Agreement between metabolite measurements available on both the MS and NMR platforms was assessed using Bland–Altman analysis.^20^ Given the differing standardisation approaches, agreement was evaluated in terms of relative concentration difference (Z-score [NMR] vs. INRT [MS]). Limits within ±0.5 SD were considered good agreement, consistent with typical technical platform variation.^21^

### Statistical analysis - weight loss trial intervention effect

To estimate the effect of bariatric surgery on metabolite levels, we applied a linear mixed-effect model (*lmer()* function from the “lmerTest” R package) where subject ID was included as a random effect to account for repeated measures (Model 1). In the below formula *M_i,t_*represents the normalised level of metabolite *i* at timepoint *t*; *timepoint_t_* is either baseline (before surgery) or at follow up (after surgery). Given this study was a single-arm cohort, the timepoint estimate reflects the change in metabolite over time following bariatric surgery. The *sub-study* term was included to account for the participants who were non-randomly allocated surgery type and in whom follow-up samples were obtained at 12-months post intervention, rather than 36-months post randomisation as in the main cohort. In addition, estimates were adjusted for hospital site, age (years), sex, and sample storage time (months).

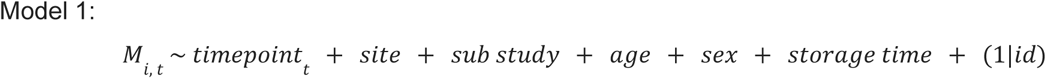

To estimate intervention effects in the two-arm DiRECT trial, we used a two-step process to capitalise on the RCT study design yet maintain comparability to the bariatric surgery study estimates. We again utilised linear mixed-effects models.

In the first step (Model 2a), each metabolite was modeled as a function of timepoint *t* (baseline or follow up), treatment allocation (intervention or control), interaction or change over time by allocation (timepoint x allocation), age at baseline (years), sex, sample storage time (months) and the minimisation variables study site (Tyneside or Scotland) and practice list size (>5700 or ≤5700). Subject ID was included as a random effect to account for repeated measures. The primary statistic of interest was the timepoint-by-allocation interaction term and its associated P-value, identifying metabolites that changed differentially over time between the dietary intervention and control groups.^22^

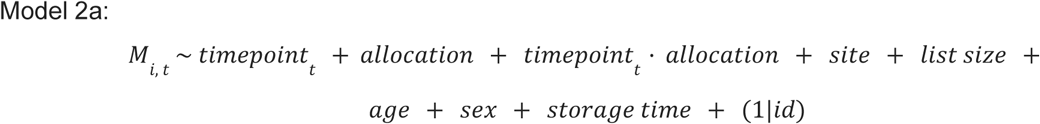

In the second step (Model 2b), to enable direct comparison with the single-arm bariatric surgery estimates, we restricted analyses to individuals in the dietary intervention arm and re-estimated timepoint effects using the same model structure as Model 1.

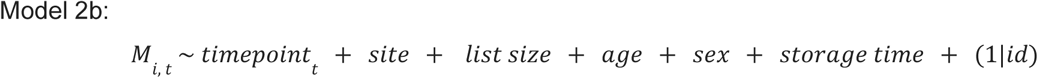

In summary, for the dietary intervention trial, P-values from the interaction term in Model 2a were used to identify metabolites significantly associated with the intervention, while timepoint effect estimates from Model 2b were used for direct comparison with the surgical trial results derived from Model 1.

### Mendelian randomization

#### Genome-wide association study data sources

The genome-wide association study (GWAS) data sources are detailed in **Supplemental Table 1**. A polygenic genetic instrument for BMI was derived from genome-wide association summary statistics from a Genetic Investigation of Anthropometric Traits (GIANT) consortium meta-analysis of over 700,000 individuals of predominantly European ancestry.^23^ GWAS summary statistics from metabolites on the NMR and MS platforms were obtained from the studies by Karjalainen et al. and Chen et al., respectively.^24,25^ To explore the associations between metabolites and HF subtypes, GWAS summary statistics for all-cause HF, non-ischaemic HF with reduced ejection fraction (HFrEF), and non-ischaemic HF with preserved ejection fraction (HFpEF) were obtained from the HERMES consortium.^26^

### Selection of instruments

Variants for inclusion in either the BMI instrument, instruments for circulating metabolites or HF outcomes, were selected based on a genome wide association study p-value threshold for the trait of P < 5 x 10^-8^. Variants were pruned to account for linkage disequilibrium using PLINK 2.0 (r^2^ < 0.001, 250 kb) with a reference panel consisting of 10,000 unrelated European ancestry participants from the UK Biobank.^27,28^ Attempting to construct genetic instruments for circulating metabolite levels has well described challenges;^25^ not least due to high correlation between individual metabolites and shared genetic aetiology. Variants selected to instrument a particular metabolite may also be associated with other, often multiple, metabolites, due to either vertical or horizontal pleiotropy, the latter of which can be problematic for MR analyses. To assess this and inform our analyses, we calculated the proportion of overlapping variants for each metabolite genetic instrument *G_i_*, with all other metabolite instruments *G_j_* (pairwise). For each pair (*G_i_*, *G_j_*), the proportion of shared variants was calculated as:

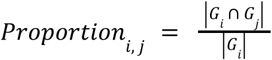

### Mendelian randomization analysis

MR was performed using a two-sample framework and the *MendelianRandomization* R package.^29^ The primary analysis used an inverse variance weighted (IVW) method, with sensitivity analyses conducted using MR Egger, weighted median and weighted mode methods to help account for pleiotropy and imperfect instruments. For the MR analysis of BMI on metabolites, MR estimates represent the change in metabolite levels (in standard deviation units) per 1 SD increase in BMI. For the MR analysis of metabolite levels on heart failure outcomes, MR estimates represent the change in log odds ratio (OR) for the outcome per 1 SD (rank normalised SD for MS metabolites; raw SD for NMR metabolites) increase in metabolite level.

### Metabolomic signature of weight loss

To distinguish metabolite changes associated with BMI from intervention-specific effects we first assessed the consistency of effect between our two primary sources of evidence - dietary intervention and bariatric surgery. Our interpretation of this overlap is that the most parsimonious explanation of shared effects is that they are driven by the most prominent joint outcome - intentional weight loss - which is also manifest as differences in BMI. Consistent effects were defined as metabolites demonstrating the same direction of association across interventions and those that reach a P value threshold of < 0.05 after Benjamini & Hochberg false discovery rate (FDR) correction.^30^ Applied within available datasets (i.e., separately for NMR and MS data), the Benjamini–Hochberg procedure controls the expected proportion of false discoveries but is less stringent than methods that control the family-wise error rate (Bonferroni or Holm). For the single-arm bariatric surgery study the P-value was drawn from the timepoint beta whereas in the calorie restriction trial the P-value was drawn from the timepoint:allocation interaction term beta. This hybrid approach incorporates information from the control arm of the dietary intervention RCT into the selection process, whilst maintaining comparable intervention estimates.

As a third line of evidence, we assessed whether metabolites showing concordant associations across both weight-loss interventions also demonstrated consistent associations with BMI at the population level. To achieve this, we performed two-sample MR analyses, using genetic variants strongly associated with BMI as instruments to proxy lifetime BMI exposure, and metabolite genetic associations as outcomes. BMI genetic variants were selected using the instrument selection procedure described above. To enable comparison of intervention-derived and MR-derived BMI estimates, weight loss associations were scaled by the average BMI change by intervention (a negative value), yielding effects per SD *increase* in BMI. Metabolites with concordant signals across both interventions and the MR analyses were prioritised for downstream assessment against heart failure subtypes (**Figure 1**).

## Results

### Study characteristics

The baseline characteristics for participants enrolled in DiRECT and By-Band-Sleeve, for which metabolite profiling was available, are presented in **Table 1**. With dietary intervention, at 12 months, mean body weight had fallen by 11.1 kg in the intervention group and by 1.4 kg in the control group (intervention associated BMI reduction of 3.7 kg/m^2^). With bariatric surgery, over median follow up of 36 (IQR 13-37) months, mean body weight fell by 29.3 kg, a BMI reduction of 10.5 kg/m^2^.

**Table 1.** Weight loss trial characteristics.

| Characteristic | Structured diet programme (DiRECT) |  |  |  | Bariatric surgery (By-Band-Sleeve)<sup>^</sup> |  |  |  |
| --- | --- | --- | --- | --- | --- | --- | --- | --- |
|  | Control arm |  | Intervention arm |  | Main (randomised) study |  | Non-randomised study |  |
| Timepoint | Baseline<br>(n = 147) | Follow-up<br>1-year<br>(n = 146) | Baseline<br>(n = 142) | Follow-up<br>1-year<br>(n = 122) | Baseline<br>(n = 969) | Follow-up<br>3-years<br>(n = 369) | Baseline<br>(n = 44) | Follow-up<br>1-year<br>(n = 44) |
| Age, years (SD) | 56.0 (7.2) | 56.2 (6.9) | 52.8 (7.6) | 53.8 (7.1) | 47 (10.8) | 49 (10.1) | 45 (10.4) | 45 (10.4) |
| Sex, male (%) | 92 (63) | 90 (62) | 80 (56) | 70 (57) | 250 (26) | 98 (27) | 6 (14) | 6 (14) |
| Weight, kg (SD) | 99.2 (15.9) | 97.8 (16.4) | 101.1 (17) | 90 (16.8) | 130.6 (23.4) | 102.1 (23.8) | 125.2 (21.0) | 93.0 (22.5) |
| BMI, kg/m<sup>2</sup> (SD) | 34.3 (4.2) | 33.9 (4.5) | 35.0 (4.5) | 31.3 (5.0) | 46.6 (6.7) | 36.3 (7.3) | 45.4 (5.8) | 33.6 (6.5) |

SD, standard deviation; kg, kilograms; m, metres
*\*numbers may differ from the published trials as these data relate to the individuals with metabolomic data passing quality control checks.*
*<sup>^</sup>all types of bariatric surgery from the By-Band-Sleeve were combined into one cohort and analysed together. There was no control arm and all individuals included underwent surgery.*

### Metabolite profiles associated with body mass index

The serum levels of 785 metabolites out of 1202 (65%) were different following bariatric surgery after FDR adjustment using the Benjamini–Hochberg procedure (FDR < 0.05). 310 demonstrated higher levels and 475 lower levels after intervention (**Supplemental Table 2**). Dietary intervention was associated with alterations in 485 of 1152 metabolites (42%) at an FDR < 0.05, with 246 demonstrating higher levels and 239 lower levels (**Supplemental Table 3**).

Of the 1422 metabolites that passed QC in either study, 932 (66%) were measured in both weight loss intervention studies and were available for comparison. The inter-study correlation between weight loss intervention effects was modest, being slightly higher for metabolites on the NMR platform compared to the MS platform - Pearson’s R 0.62 (95%CI 0.53-0.70) vs. 0.53 (95%CI 0.47-0.58), respectively (**Supplemental Figure 3**).

Thirty-three metabolites feature in both the MS and MNR platforms and were available for comparison within studies. The intra-study correlation between these measures was high in both weight loss trials: Pearson’s R 0.98 (95%CI 0.94-0.99) (bariatric surgery), and 0.90 (95%CI 0.73-0.96) (dietary intervention) (**Supplemental Figure 4**). In the Bland-Altman analysis the mean difference (MS - NMR) was 0.010, within 95% limits of agreement ranging from −0.29 to 0.31. One metabolite (3%, glutamine) fell outside the 95% limits of agreement, less than the expected 5% if differences were normally distributed - indicating excellent agreement between platforms (**Supplemental Figure 5**).

For 240 metabolites there was both strong evidence of association with intervention and consistent effect direction across both studies (**Figure 2 & Supplemental Table 4**). Of these, 112 (47%) increased with weight loss intervention and 128 (53%) decreased. The metabolites with the largest intervention-associated level increases included 1-oleoyl-GPC (18:1), 1-(1-enyl-palmitoyl)-2-oleoyl-GPC (P-16:0/18:1), and 1-methylxanthine. The metabolites with the largest intervention-associated level decreases included valine, Behenoyl dihydrosphingomyelin (d18:0/22:0), and phospholipids to total lipids ratio in chylomicrons and extremely large VLDL.

**Figure 2.**
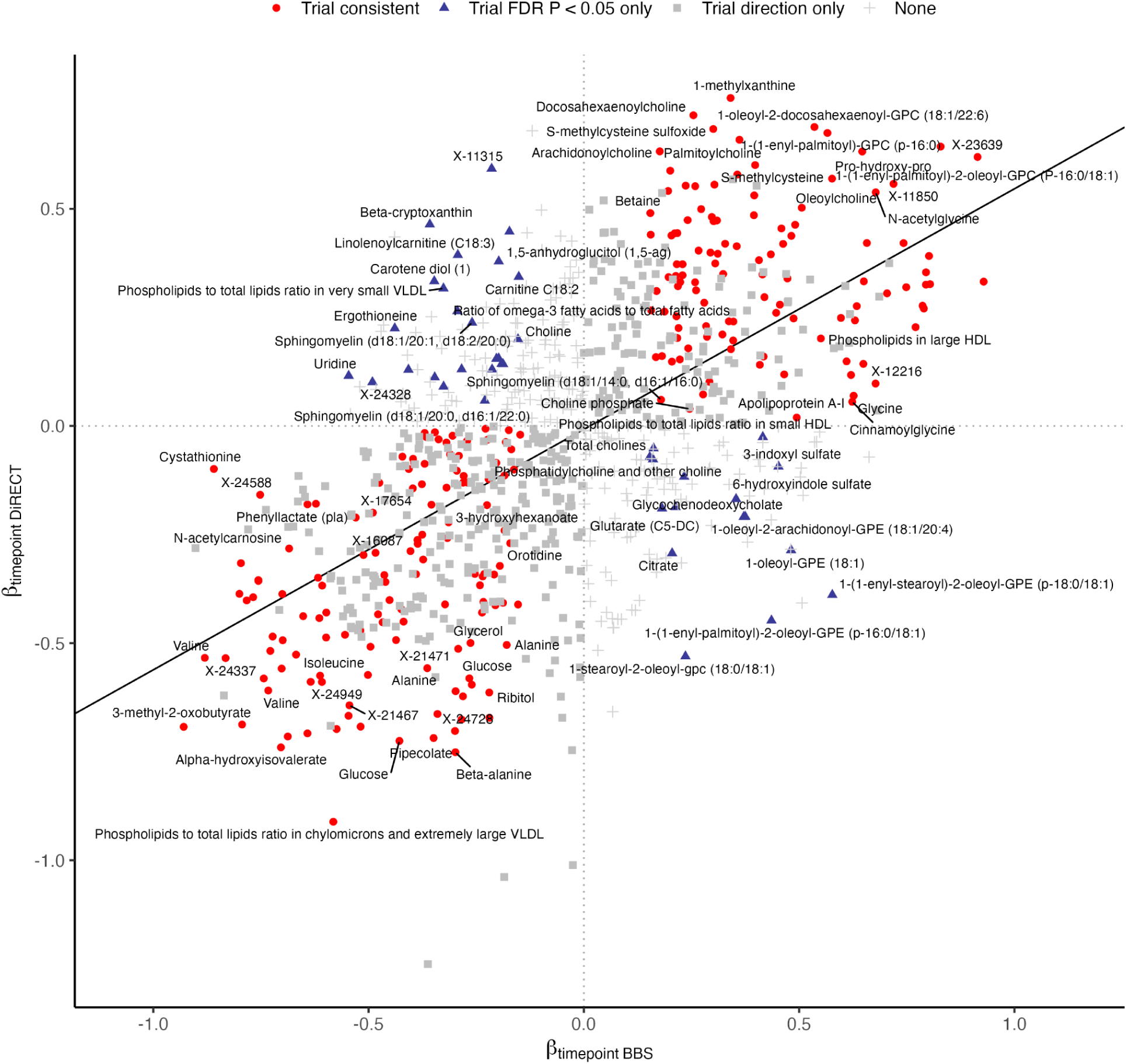
Comparison of estimates for the effect of weight loss intervention on metabolites. Scatterplot comparing estimated effects of bariatric surgery (x-axis) and calorie restriction (y-axis) on circulating metabolite levels. Each point represents a metabolite. Non-overlapping metabolites names are labelled. Red circles indicate metabolites with consistent directions of effect and FDR-significant associations in both interventions; blue triangles indicate metabolites with opposite directions of effect and FDR-significant associations; grey squares denote metabolites with concordant directions but not FDR-significant; and grey plus signs indicate metabolites without significant association or direction in either study.

The MR estimates for the difference in metabolite levels per 1 SD (∼ 4.8 kg/m^2^) higher on average life course BMI are presented in **Supplemental Table 4**. The independent genetic variants used to instrument BMI are present in **Supplemental Table 5**. To harmonise weight loss intervention estimates to the MR estimates, intervention association statistics were converted to per SD BMI change effects by dividing by the trial average SD BMI change (a negative value resulting in sign inversion). Among the 240 metabolites demonstrating consistent weight loss effects across both interventions, 229 (95%) had corresponding lifetime BMI MR estimates. Of these, 153 (67%) exhibited concordant directions of association at an FDR-adjusted P value < 0.05. The strongest evidence for lifetime BMI increasing effect was for 5-methylthioadenosine, whereas the strongest evidence for a BMI decreasing effect was with the metabolite 1-(1-enyl-palmitoyl)-2-oleoyl-GPC (P-16:0/18:1). Metabolites from lipid (31%), lipoprotein (26%), and amino-acids (20%) biochemical classes formed the majority of the consistent signals seen across all three lines of evidence. These 153 concordant effect metabolites are hereafter referred to as ‘BMI-metabolites’ (**Figure 3**).

**Figure 3.**
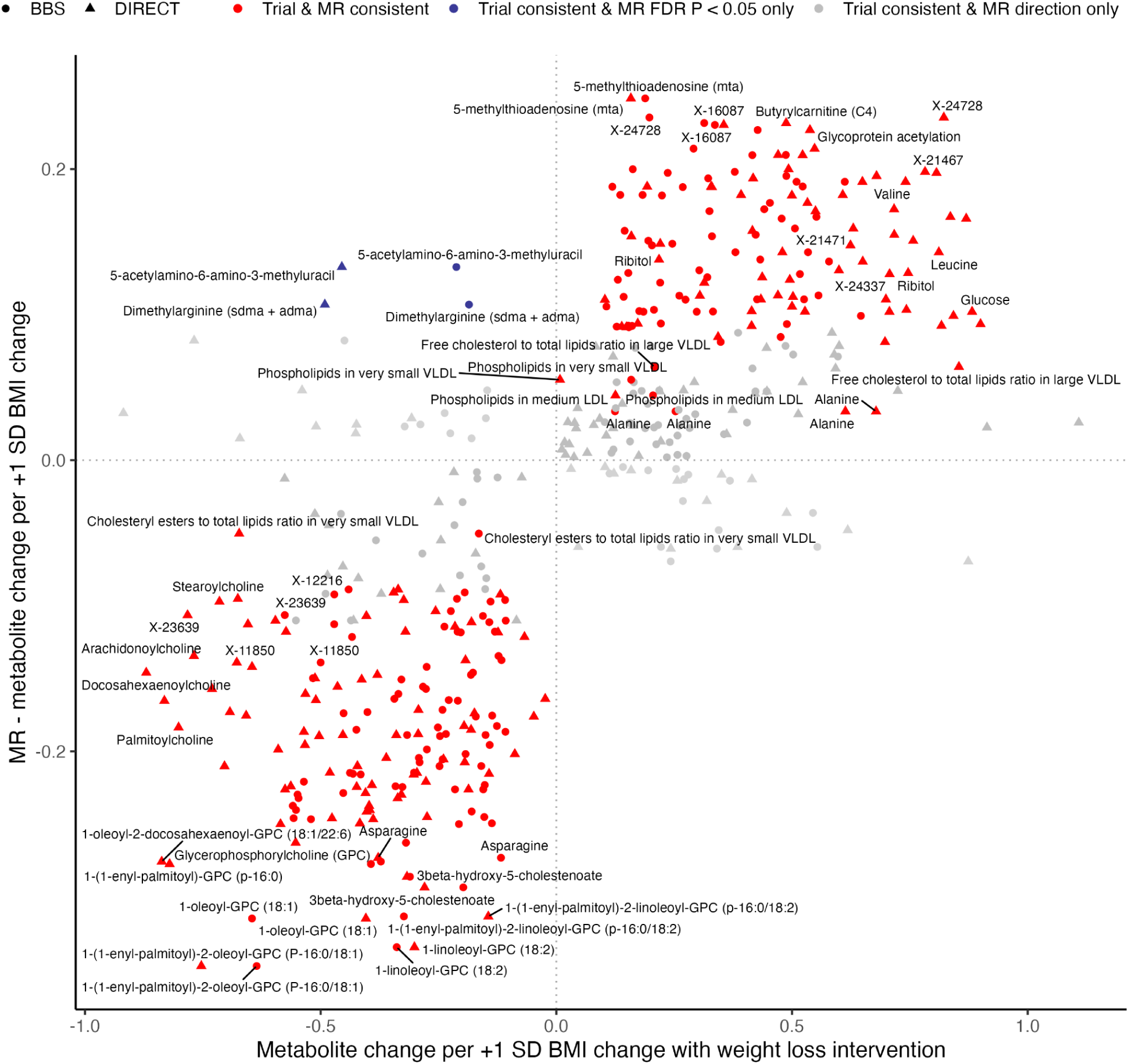
Comparison of estimates for the effect of weight loss intervention and life-time body mass index exposure on metabolites. Comparison of metabolite effect estimates from BMI MR analyses (x-axis) and weight-loss interventions (y-axis), where circles represent bariatric surgery effects and triangles represent calorie restriction effects. Red points indicate metabolites with consistent directions of effect across both interventions and MR, all FDR-significant. Blue points indicate metabolites with concordant intervention effects but opposite-direction, FDR-significant MR associations. Dark grey points denote metabolites with consistent directions across interventions and MR but without FDR-significant MR associations. Light grey points indicate metabolites with consistent intervention effects but neither significant nor directionally consistent MR results.

### BMI-metabolite associations with heart failure

To identify metabolic factors underlying the beneficial effects of weight loss on HF, we investigated the association of BMI-metabolites with heart failure and its subtypes, using Mendelian randomization.

Genetic instruments were available given the prespecified parameters for 133 of 153 (87%) BMI-metabolites, with instrument sizes (number of variants) ranging from 1 to 161. BMI-metabolite instruments demonstrated a wide range of specificity, with the percentage of shared variants with any other metabolite (BMI-metabolites or otherwise) instrument ranging from 0-100% (**Supplemental Table 6**). Of all variants selected into at least one BMI-metabolite instrument, 54% were also genome wide hits for ‘non-BMI’ metabolites (**Supplemental Figure 6**). Instruments for metabolites from the lipoprotein and lipid classes were notably non-specific, overlapping largely with metabolites from the same class. For example, total lipids in large high-density lipoproteins (HDL) was instrumented with 135 variants, however shared 99% of these with the instrument for concentration of large HDL particles and 29% with Apolipoprotein A-I, whereas there was much less overlap with molecules of other classes, such as creatinine (1%) or glutamine (1%) (**Supplemental Table 6 & Supplemental Figure 7**).

In the primary IVW-MR analysis, 44 BMI-metabolites were associated with HF subtypes at an unadjusted P value < 0.05 (**Figure 4** & **Supplemental Table 8**). These were predominantly lipid and lipoprotein metabolites for the outcome of all-cause HF, a phenotype that includes a high proportion of cases with atherosclerotic / ischaemic aetiology. Six amino acid BMI-metabolite traits associated with all-cause HF: glutamine, N2,N5-diacetylornithine, 4-hydroxyphenylpyruvate, 5-oxoproline, N-acetyleglycine and 2-hydroxybutyrate/2-hydroxyisobutyrate levels, as well as the carbohydrate BMI-metabolite ribitol. Lipid and lipoprotein traits were under-represented for the outcome of non-ischaemic HFpEF, compared to all-cause HF. Instead, three amino acids - N2,N5-diacetylornithine, gamma-glutamylvaline and asparagine - and three co-factors or xenobiotics - oxalate, tartronate, and threonate - were associated with the outcome at an unadjusted P value < 0.05. For non-ischaemic HFrEF, several low density lipoprotein (LDL) traits displayed an inverse association with the outcome, as well as similar associations with oxalate, tartronate, and threonate.

**Figure 4.**
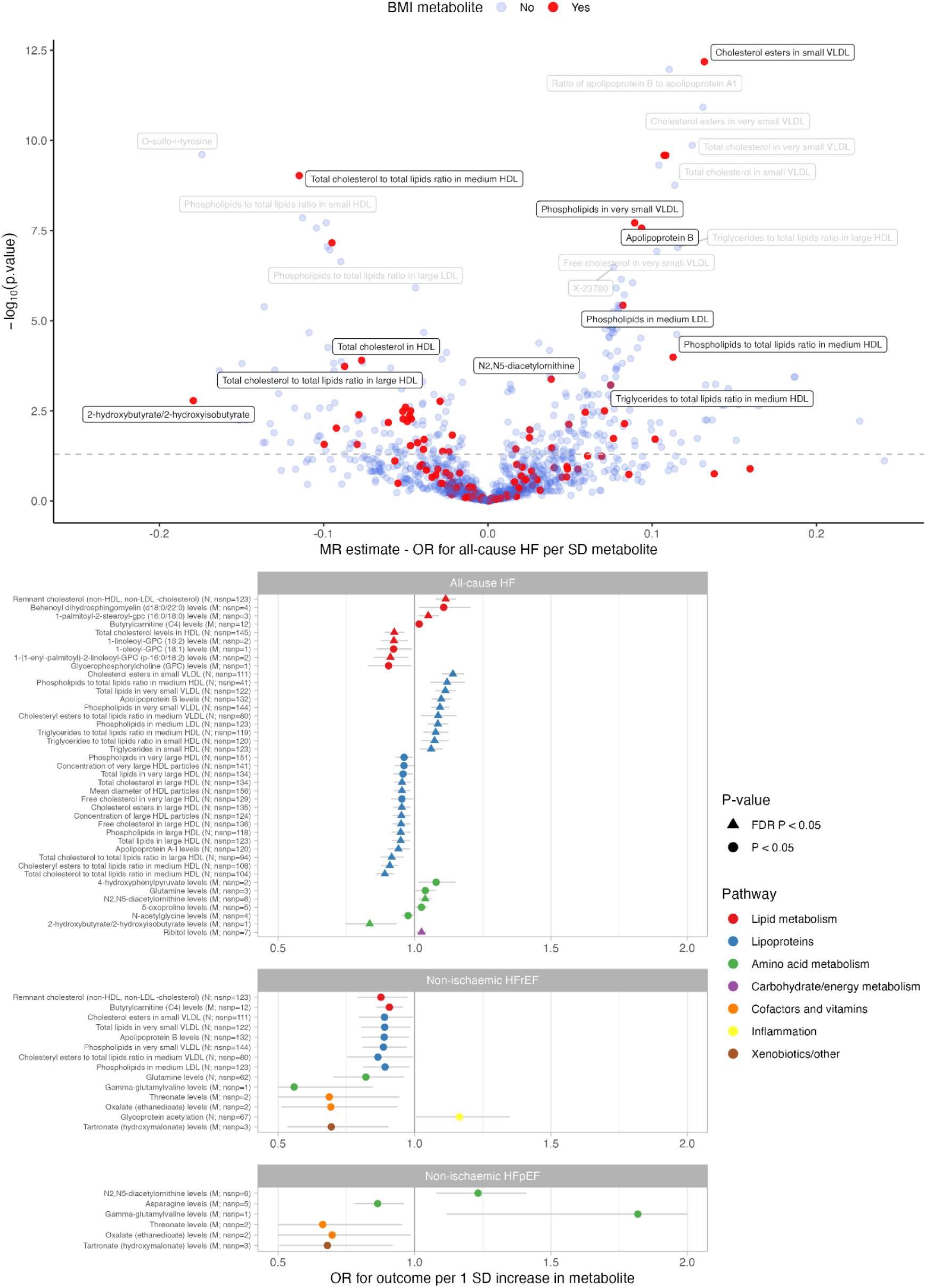
Mendelian randomization estimates for the effect of metabolite levels on heart failure outcomes. The top panel shows a volcano plot of inverse variance weighted Mendelian randomisation estimates for the effect of metabolite levels and all-cause heart failure, with effect estimates plotted against –log₁₀(P-value). The bottom panel presents a stacked forest plot summarising metabolites with nominal (P < 0.05; circles) or FDR-significant (FDR < 0.05; triangles) MR associations, grouped by metabolic pathway (indicated by colour). Results are subdivided by heart failure subtype: all-cause HF, HFrEF, and HFpEF. Error bars represent the 95% confidence interval.

### Exploratory post-hoc analyses

To further contextualise our results, first, we compared the overlap of our BMI-metabolites with previously published observational associations with incident HF: MS metabolites measured in 3,719 participants of the ARIC study,^31^ and NMR metabolites measured in ∼500,000 individuals enrolled in the UK Biobank.^32,33^ For the MS metabolites hazard ratios had been derived for incidence all-cause HF, HFpEF, and HFrEF, in a Cox regression model adjusted for clinical risk factors including age, sex, ethnicity, center, heart rate, BMI, smoking status, systolic blood pressure, blood pressure medication use, prevalent diabetes, prevalent coronary heart disease, and prevalent atrial fibrillation (described as model 1 in their report).^31^ For the NMR metabolites hazard ratios were derived for all-cause HF outcome only, in a Cox regression model adjusted for age, sex and UK Biobank assessment centre.

Sixty-nine of the 153 BMI-metabolites (45%) also demonstrated an observational association with incident all-cause HF at an FDR-adjusted P < 0.05. For HFpEF and HFrEF, observational data were only available for metabolites measured on the MS platform. Of the 101 MS BMI-metabolites, 13 were also associated with HFpEF, four of which were unique to HFpEF and nine which were also associated with all-cause HF. No BMI-metabolites overlapped with those associated with HFrEF (**Supplemental Figure 8 & Supplemental Table 8**).

Next, we looked for consistency of BMI-metabolite effect in the observational data and our primary IVW MR analysis of the effect of metabolites on HF outcomes (**Supplemental Figure 9**).

For all-cause HF, HDL- and LDL-related measures predominated among the traits associated with HF risk in both the observational and MR analyses. Total cholesterol in HDL particles was inversely associated with risk of all-cause HF in both MR (OR 0.93 per SD 95%CI 0.89-0.96, P = 1.2 x 10^-4^) and observational analyses (HR 0.76 per SD 95%CI 0.75-0.77, P = 8.0 x 10^-177^) (**Supplemental Table 8**). This trait was inversely associated with BMI in both weight-loss interventions and was inversely associated with BMI predicted using genetic data. Its genetic instrument comprised 146 variants (F stat 97) and shared variants with 251 other metabolites, mainly other HDL traits - for example, an overlap of 42% with Apolipoprotein A-I levels (**Supplemental Figure 7 & Supplemental Table 6**). Conversely, butyrylcarnitine (C4) levels were associated with higher all-cause HF risk in both the MR analysis (OR 1.02 per SD 95%CI 1.00-1.03, P = 0.04) and observational data (HR 1.18 per SD 95%CI 1.05-1.33, P = 0.025) (**Supplemental Table 8**). This trait was positively associated with BMI in both weight-loss interventions, and positively associated with BMI predicted using genetic data. Its genetic instrument comprised 12 variants (F stat 307), sharing a relatively small proportion of variants (maximum 15%) with 23 other metabolite traits, mainly other carnitine molecules (**Supplemental Figure 7 & Supplemental Table 6**). Interestingly, several VLDL traits showed discordant associations with all-cause HF between observational and MR analysis. Total lipids, phospholipids, and cholesterol ester VLDL content, as well as apolipoprotein B levels, all decreased following weight loss intervention and were inversely associated with life-time BMI (**Supplemental Figure 9 & Supplemental Table 8**). In the MR analyses, these traits were positively associated with all-cause HF risk, whereas the published observational estimates indicated inverse associations. However, these observational estimates were largely explained by widespread use of lipid-lowering medications and attenuated or reversed when individuals on lipid-lowering medications were removed.^33^

Comparing the associations for MS metabolites with HFpEF, we saw increased gamma-glutamylvaline levels associated with risk of HFpEF in both the MR analysis (OR 1.82 per SD 95%CI 1.12-2.96, P = 0.016) and observational data (HR 1.42 per SD 95%CI 1.14-1.77, P = 0.015) (**Supplemental Table 8**). This BMI-metabolite was positively associated with BMI in both weight-loss interventions, and positively associated with predicted BMI using genetic data. The gamma-glutamylvaline instrument contained a single variant, limiting MR sensitivity analyses. Higher asparagine levels were associated with a decreased risk of HFpEF in our MR analysis (OR 0.86 per SD 95%CI 0.78-0.96, P = 0.007) and demonstrated consistent direction in the observational data (HR 0.79 per SD 95%CI 0.64-0.95, P = 0.055) but with borderline FDR-adjusted significance. The asparagine MR instrument was made up of 5 variants (rs715, rs10459574, rs115395195, rs28393117, rs8012505) with an F statistic of 206. Three of the variants are within 250 kb of the asparaginase and asparagine synthetase genes, increasing the plausibility of the instrument. Two variants were shared with other metabolite instruments: rs715 with glutamine degradant, gamma-glutamylhistidine, histidine, and unknown compound X-25420, and rs8012505 appeared in the instrument for the related compound N-acetylasparagine (**Supplemental Table 9**). Leave-one-out analysis demonstrated consistent effect direction (**Supplemental Figure 10**) and cis-MR of the asparaginase gene region variants (rs55798317 & rs8012505) demonstrated similar results, as well as the cis-MR result (Wald ratio) using the single variant in the asparagine synthetase gene region (rs28393117), albeit with wide confidence limits (**Figure 5**).

**Figure 5.**
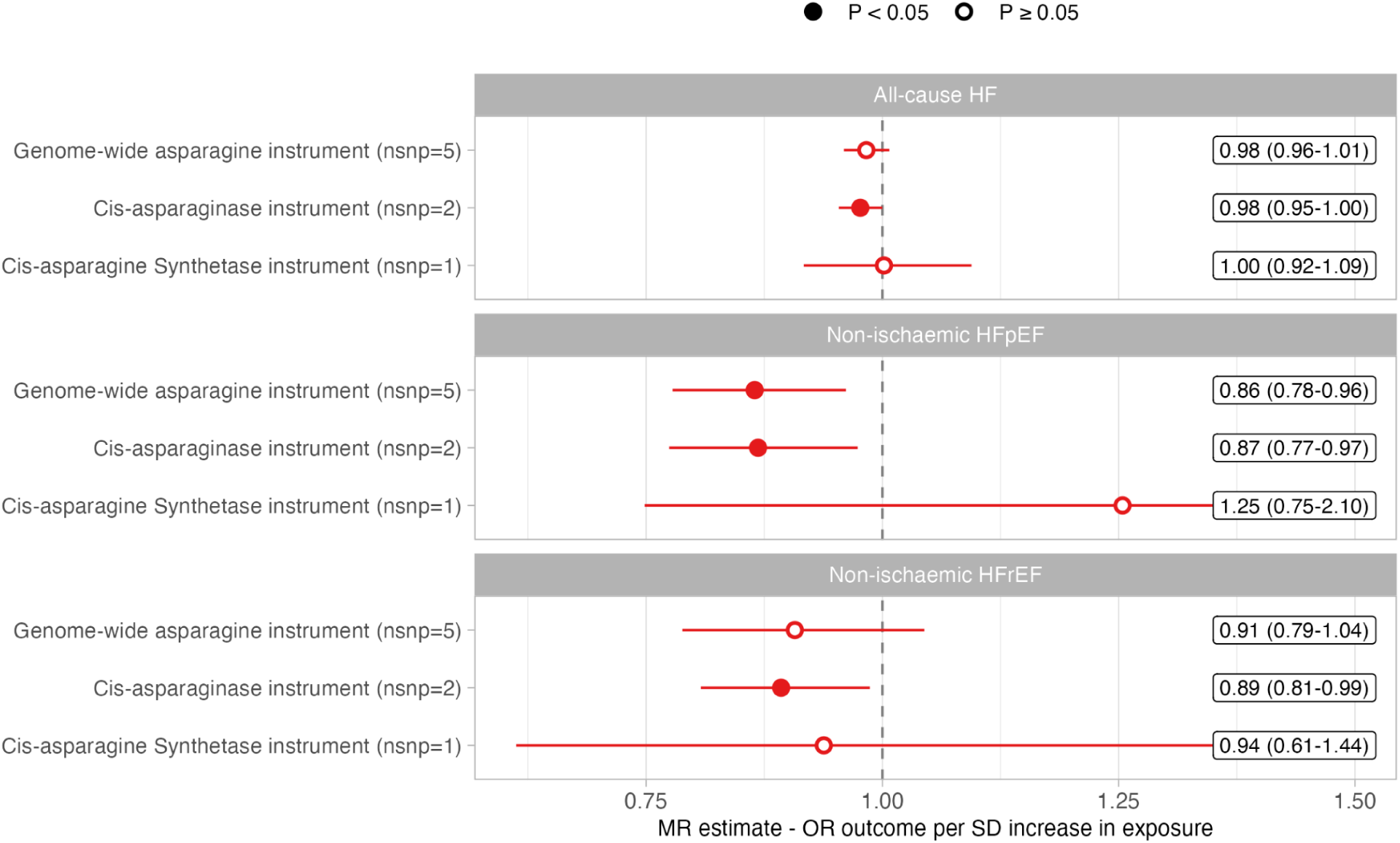
IVW Mendelian randomization estimates for the effect of asparagine levels on heart failure subtypes. Inverse-variance weighted Mendelian randomisation (MR) estimates are shown for the association between genetically predicted asparagine levels and heart failure outcomes. The x-axis represents the MR effect estimate, and the y-axis represents the genetic instrument used: *genome-wide*, *cis-asparaginase*, and *cis-asparagine synthetase variants*. Closed circles indicate associations with *P < 0.05*, and open circles indicate *P ≥ 0.05*. Error bars represent *95% confidence intervals*.

None of the BMI-metabolites were strongly associated with HFrEF in both the MR and observational analyses.

## Discussion

In this study, we integrated metabolomic profiling from different types of weight loss intervention - dietary intervention and bariatric surgery - as well as with life-time BMI exposure using Mendelian randomization. We identified 153 BMI-metabolites that were consistently altered across interventions and lifetime BMI exposure, of which 44 demonstrated potential causal links with HF and its subtypes.

Obesity is a major risk factor for the development of HF across a range of aetiologies and subtypes, including HFrEF and HFpEF. Excess adiposity is now modifiable at a patient and potentially population level, with the recent development of effective weight loss therapies, yet the contributing molecular pathways remain elusive. Understanding these pathways is important, as not all individuals are eligible for, respond to, or tolerate current weight-loss interventions. In addition, identifying downstream metabolic mediators may reveal novel therapeutic targets or biomarkers that could potentially be modulated independently of weight. Addressing the overarching need to better understand the biological implications of weight difference and living with obesity (often captured by BMI variation), we have been able to triangulate evidence from data with different sources of variation, error and bias to identify consistent metabolic signals related to a latent set of shared conditions robustly capturing weight.

Around half of the measured metabolites show evidence for changes following either bariatric surgery or dietary intervention, with lipids, lipoproteins and several amino acids the main responders. There are well known amino acid changes with bariatric surgery that are replicated in our data, with weight loss associated with reductions in valine, leucine, and isoleucine and increases in glycine levels.^34^

Moderate correlation between the effects of the contrasting interventions suggests both shared and intervention-specific effects of the circulating metabolome. The contrasting study designs introduce different sources of bias, however, this is informative as these biases are likely different and therefore shared effects across studies consistent with adiposity-specific signals. This does not rule out common alternative explanations, such as engagement with other lifestyle choices by virtue of being enrolled in a clinical trial, or medications common to weight loss programmes. Nearly all of the metabolites with consistent BMI effects between weight-loss interventions also showed consistent associations with BMI predicted using genetic data. This convergence provides a natural experiment in gene-environment equivalence, whereby genetic proxies and intervention-induced changes point toward the same underlying biology. Further metabolic profiling in other weight-loss interventions, such as incretin-based therapies, will be needed to refine the common signals of adiposity from treatment-specific effects.

Weight loss in individuals with obesity is associated with many benefits and whilst historically this has been difficult to achieve and maintain, there are a growing number of efficacious intervention strategies - including dietary interventions, surgery and pharmaceutical agents. However, the underlying mechanisms by which weight loss inducing strategies translate into improved outcomes is incompletely understood. For example, it is not known whether the benefits are driven solely by weight loss itself or whether direct intervention effects independent of anthropometric change also contribute - a relationship that likely varies across specific diseases and outcomes.^35^ The resultant - and often unseen heterogeneity - is important as there now exists clinical choice with respect to weight-loss interventions and currently limited basal biological evidence able to discriminate between them. In addition, greater understanding of the underlying biological pathways should aid in further drug development efforts to either induce weight loss or mimic all or part of its beneficial effect. Specific to HF, emerging trial evidence supports the role of weight loss to improve outcomes in individuals with established HFpEF,^36,37^ but there is concern regarding weight loss strategies in HF with signals for harm in reduced ejection fraction (HFrEF) from small RCTs as well as observational data.^38–40^

Our MR analysis of BMI-associated metabolites revealed distinct associations with HF subtypes. Lipid and lipoprotein metabolites were strongly linked to all-cause HF, consistent with the phenotype given that nearly a third of cases had a history of myocardial infarction.^26^ Aside from lipoprotein traits there were potential causal signals for less well established lipid molecules. Butyrylcarnitine (C4) levels decrease with weight loss, increase with lifetime BMI and were associated with a 1.02 (1.00-1.03, P = 0.036) increased odds of all-cause HF in this analysis. Butyrylcarnitine (C4) has also been shown to have prognostic value in HF patients, predicting HF hospitalisation and death.^41,42^ Acetylcarnitines are products of mitochondrial beta-oxidation and in HF patients correlate with Brain Natriuretic Peptide (BNP) levels.^42^ Accumulation is potentially indicative of the mitochondrial dysfunction seen in HF, where anomalous lipid and energy metabolism, through defective beta-oxidation of medium and short-chain fatty acids or impaired coupling between fatty acid oxidation and electron transport chain, leading to increased levels.^43,44^ The butyrylcarnitine instrument shared variants with 23 other metabolites, mainly within the carnitine metabolism network.

Lipoprotein and lipid metabolism plays a well known role in the HF aetiology, however, a substantial proportion of the risk of BMI in the development of HF is known to remain even after controlling for such metabolic risk factors.^45,46^ Genetic analyses estimate that only 40% of the risk is mediated through coronary heart disease, hypertension, diabetes, and atrial fibrillation,^47^ and even in HF of non-ischaemic aetiology BMI remains one of the strongest predictors.^26^ Whilst the number of non-lipid/lipoprotein metabolites associated with BMI across multiple sources of BMI variation is relatively small, our data suggest a role for several non-lipid components of the adiposity metabolic signature in transducing excess adiposity through to HF risk.

In these data N-acetylglycine levels increased with weight loss, decreased with lifetime BMI exposure, and were inversely associated with all-cause HF risk in our MR analysis (OR 0.98; 95%CI 0.95-1.00). The genetic instrument comprised four variants but showed overlap with 53 other metabolite traits, predominantly other acylglycines, N-acetylated amino acids, and glycine-related metabolites, along with several lipoprotein and amino acid measures. Interestingly, others have reported genetic analyses showing that higher GLP1R activity may be causally associated with N-acetylglycine levels and that the metabolite may mediate ∼8% of the total protective effect of GLPR agonism on heart failure risk.^48^ In addition, as previously reported, our dietary intervention is associated with significant reductions in blood pressure,^49^ which may in part be mediated by this metabolite, as other MR analyses suggest a potential causal effect of higher N-acetylglycine levels on lower diastolic blood pressure.^50^

Our analyses highlight a potential role protective role for LDL traits in non-ischaemic HFrEF, this controversial relationship has been reported previously yet the underlying factors driving this association remain unclear.^51,52^ In addition, we found links between non-ischaemic HF and several amino acids, cofactor molecules and xenobiotics. Asparagine levels increase with weight loss through both dietary intervention and bariatric surgery and are negatively associated with lifetime BMI. There is a close relationship between dysregulated amino acids and diseases such as HF,^53^ and our MR analyses suggest asparagine metabolic pathways may be involved in non-ischaemic HF pathogenesis, with higher circulating asparagine associated with reduced odds of HFpEF. Asparagine synthetase (ASNS), the enzyme responsible for the production of asparagine, has been shown to regulate mouse myocardial cell differentiation, although in the opposite direction to our finding. Mouse ASNS knockdown resulted in reduced neonatal cardiomyocyte asparagine levels, yet an increase in cardiomyocyte death and reduced regeneration in response to myocardial infarction.^54^

Oxalate (ethanedioate) and tartronate (hydroxymalonate) levels increase with weight loss and decrease with lifetime BMI yet in our MR analyses these two metabolites were associated with around a 30% reduced risk of both non-ischaemic HFpEF and HFrEF (OR 0.69; 95%CI 0.51-0.94, OR 0.70; 95%CI 0.49-0.99, respectively). Genetic instruments for both traits comprised only a small number of variants: the two-variant oxalate instrument overlapped with the non-BMI metabolites threonate (rs62381198) and glycerate (rs62381616), while the three-variant tartronate instrument shared one variant (rs62381620) with that for threonate levels. Elevated serum oxalate is known to be associated with adverse cardiovascular outcomes, particularly in individuals with kidney failure.^55,56^ However, oxalate and threonate are both degradation products of ascorbic acid metabolism and may be markers of plant foods rich in vitamin C or green vegetable intake,^57–59^ suggesting that these discordant signals may reflect a complex interplay between disease states and potential dietary changes in our interventions.

It is unlikely that all pathway perturbations associated with excess adiposity are relevant in the development of all adiposity-linked diseases. Attempting to define the pathways relevant to specific diseases is a worthy endeavor that may lead to condition-specific treatments outside simply inducing weight loss, though admittedly intentional weight loss brings many additional benefits. In addition, defining a reproducible metabolic signature of weight loss could have practical implications in managing and monitoring clinical trials. If a consistent metabolic signature can be defined and shown to be associated with improved clinical outcomes, it could serve as an intermediate biomarker in early-phase drug trials, helping accelerate the validation of new weight loss therapeutics.

There are several important limitations to the data presented. We bring together data from two different weight-loss interventions - the DiRECT trial enrolled individuals with obesity and type 2 diabetes whereas the By-Band-Sleeve trial enrolled a broader population of individuals with obesity. The dietary intervention trial was substantially smaller than the bariatric surgery study with a consequent reduction in power. In our analysis this will have resulted in some adiposity-related metabolites being filtered out due to apparent weak association. Incorporation of other weight-loss interventions, such as individuals treated with incretin-based therapies, as well as larger sample sizes, will aid in further confirming the metabolite signature of weight loss versus intervention-specific effects.^35^ Instrumentation of specific metabolites is challenging owing to the non-specific nature of many of the instruments, in this regard our MR results should not be interpreted as evidence that any particular metabolite is directly causal for a disease outcome, rather as evidence of potential pathway involvement and a basis for further investigation as to a metabolite’s specific role. In our exploratory comparative analysis, we describe previously published observational estimates for the association of MS metabolites and incident HFrEF and HFpEF. However, these data include individuals with both non-ischaemic and ischaemic HF, whereas our genetic analyses were restricted to non-ischaemic HFrEF and HFpEF. As such, the comparisons between the genetic and observational findings across these HF subgroups should be interpreted cautiously and considered hypothesis-generating.

In conclusion, we have characterised a robust metabolic signature of adiposity across multiple independent sources of BMI variation. This integrative approach disentangles shared adiposity-related biological signals from intervention-specific and non-adiposity signals captured through BMI, thus isolating metabolic pathways likely to be downstream of adiposity and upstream of HF risk. These findings highlight metabolic components of adverse adiposity most relevant to heart failure development and suggest potential metabolic targets for prevention and treatment. Future work will refine this further through focus on comparisons with other weight-loss interventions and pursuing consistent signals to determine the potential to replicate the beneficial effects of weight loss for HF and other obesity-related diseases through pharmacological means.

## Author Contributions

Conceptualization: NJT, LC

Data curation: NS, MLS, Graziella Mazza, Eleanor Gidman

Formal analysis: NS, MLS

Funding acquisition: NJT, LC

Methodology: NS, MLS, LP, RTL, NJT, LC

Project administration: LC, NJT

Supervision: LC, LP, RTL, NJT

Visualization: NS

Writing – original draft: NS

Writing – review & editing: all authors

## Data Availability

All data produced in the present work are contained in the manuscript

## Acknowledgements

By-Band-Sleeve: The By-Band-Sleeve trial is led by JM Blazeby (Chief Investigator) and CA Rogers (lead methodologist). We acknowledge the support of all other members of the NIHR By-Band-Sleeve Management Group: Robert C Andrews, John Bessant, James P Byrne, Nicholas Carter, Caroline Clay, Jenny L Donovan, Eleanor Gidman, Graziella Mazza, Mary O’Kane, Barnaby C Reeves, Nicki Salter, Janice L Tompson, Richard Welbourn and Sarah Wordsworth. We also thank all the By-Band-Sleeve contributors including the investigators, research dieticians and nurses, the independent trial steering committee and data monitoring and safety committee. We are grateful to all the patients who participated in this trial.

DiRECT: We thank the National Health Service (NHS) Primary Care Research Network and North East Commissioning Support for their support and valuable input to recruitment. We thank Elaine Butler, Josephine Cooney, Sara-Jane Duffus, and Philip Stewart from the University of Glasgow for providing technical assistance; Helen Pilkington from the Newcastle upon Tyne Hospitals NHS Foundation Trust for providing research nurse support; and Sarah Weeden and Sarah Cadzow from the Robertson Centre for Biostatistics. We thank Wilma Leslie and research dietitians Louise McCombie, George Thom, Naomi Brosnahan and Alison Barnes for their contributions to the collection and transport of samples. We are enormously grateful to the GP practices, health-care professionals, and volunteers for their participation.

## Data and code availability

The analysis was conducted using a Snakemake (version 7.26) workflow.^60^ The code will be made available on final publication on GitHub.

Anonymised individual patient data from the By-Band-Sleeve trial will be made available upon request to the chief investigator for secondary research, conditional on assurance from the secondary researcher that the proposed use of the data is compliant with the Medical Research Council Policy on Data Sharing regarding scientific quality, ethical requirements, and value for money, and is compliant with the National Institute for Health and Care Research policy on data sharing. A minimum requirement with respect to scientific quality will be a publicly available prespecified protocol describing the purpose, methods, and analysis of the secondary research (e.g., a protocol for a Cochrane systematic review), approved by a UK Research Ethics Committee or other similar, approved ethics review body. Participant identifiers will not be passed on to any third party.

The terms of participant consent in DiRECT do not allow making the study data freely available in its raw form. The data used for analysis will be placed on a research data repository (https://researchdata.gla.ac.uk/) with access given to researchers subject to appropriate data sharing agreements.

---

**Figure 1.**
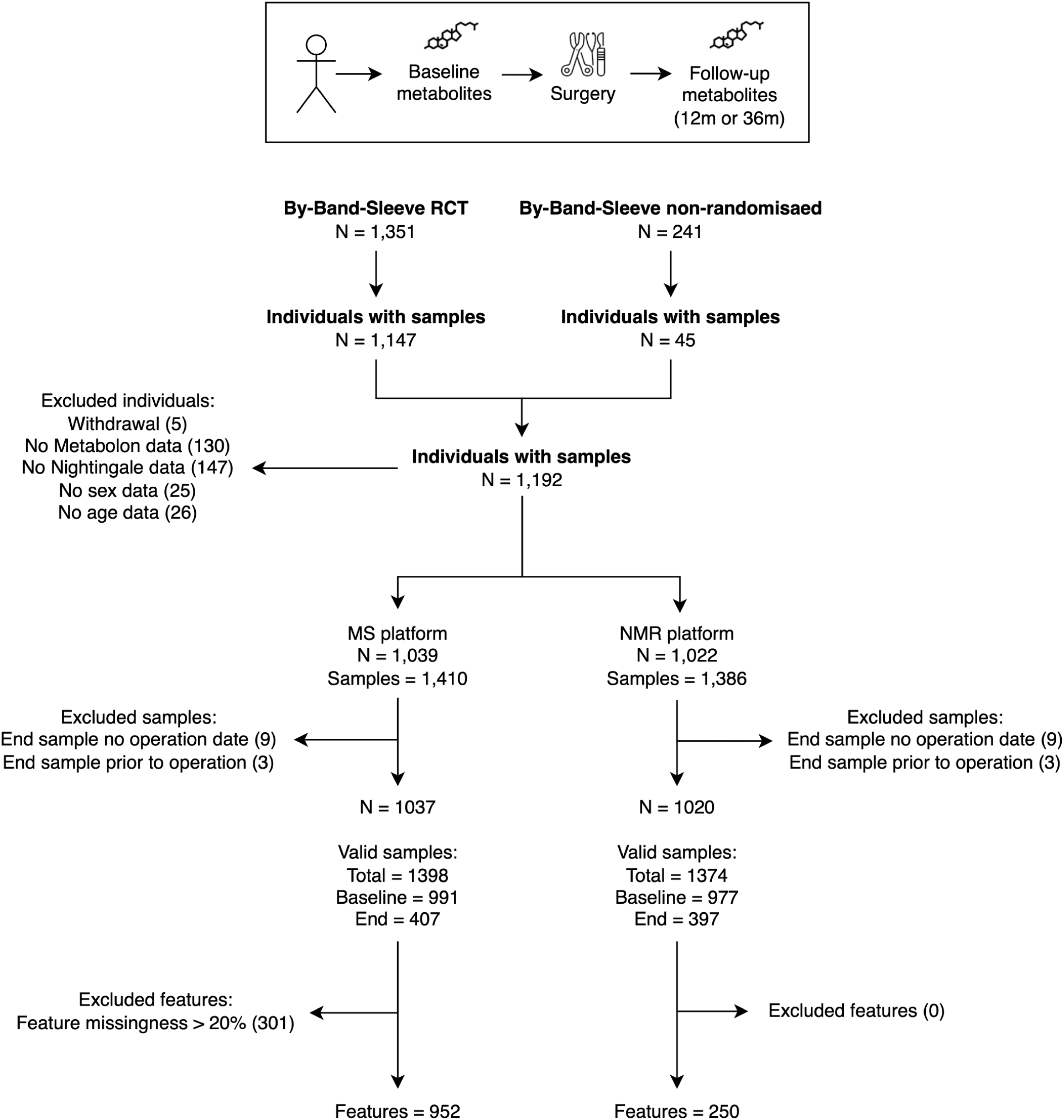
By-Band-Sleeve sample and feature flow. Study flow diagram from the By-Band-Sleeve bariatric surgery trial. The top panel depicts the single-arm trial design with all individuals undergoing bariatric surgery. The bottom panel shows the flow of individuals (N) and their samples through the quality control pipeline. Metabolites were measured on two profiling platforms - mass spectroscopy (MS) and nuclear magnetic resonance (NMR). The total number of samples available for analysis, and the split between those available at baseline and the end of follow up, are shown. The number of *features* represents the number of individual metabolites available for analysis after quality control.

**Figure 2.**
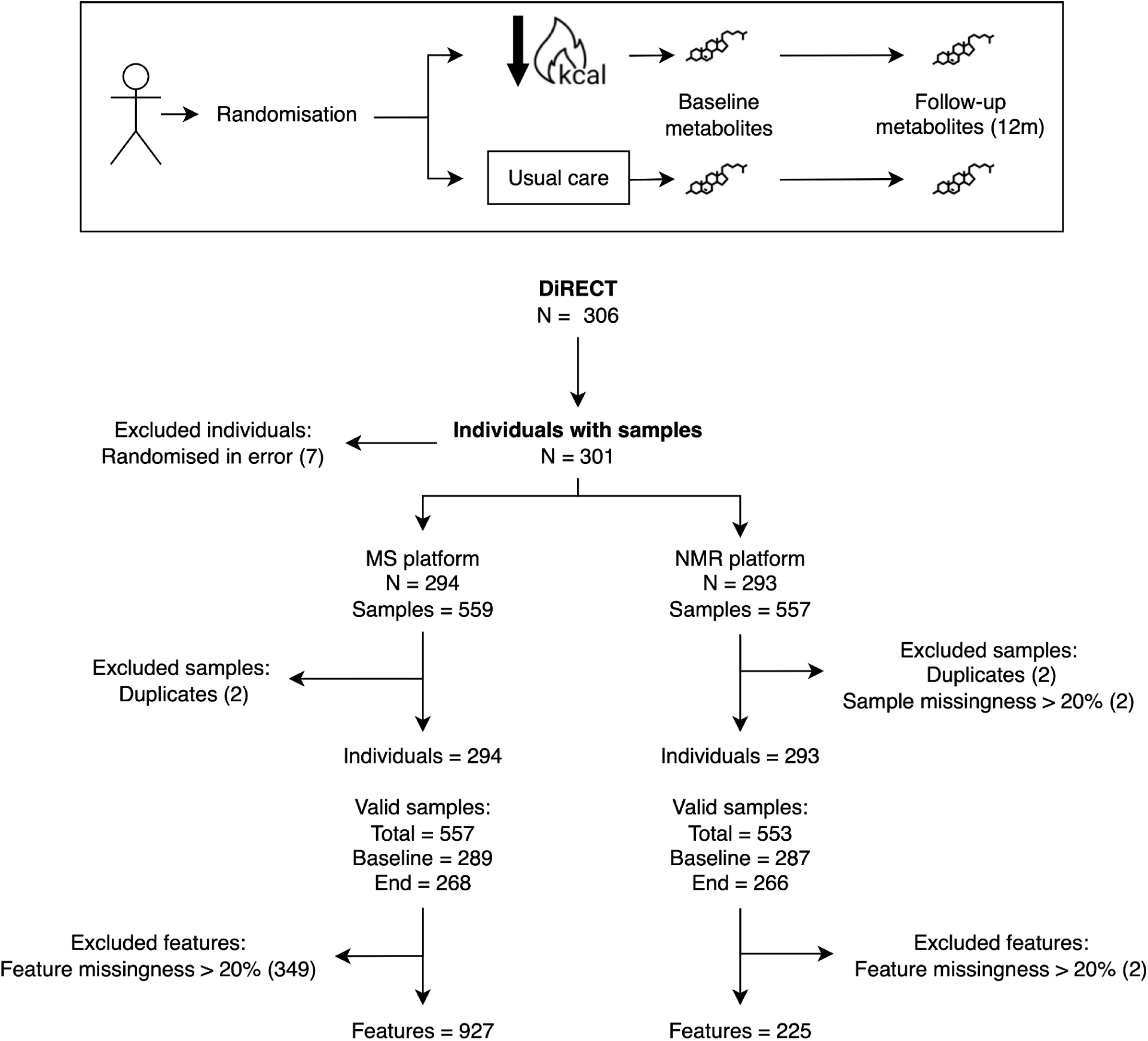
DiRECT sample and feature flow. Study flow diagram from the DiRECT dietary intervention trial. The top panel depicts the randomised control trial design with individuals allocated to either the dietary intervention or control arms. The bottom panel shows the flow of individuals (N) and their samples through the quality control pipeline. Metabolites were measured on two profiling platforms - mass spectroscopy (MS) and nuclear magnetic resonance (NMR). The total number of samples available for analysis, and the split between those available at baseline and the end of follow up, are shown. The number of *features* represents the number of individual metabolites available for analysis after quality control.

**Figure 3.**
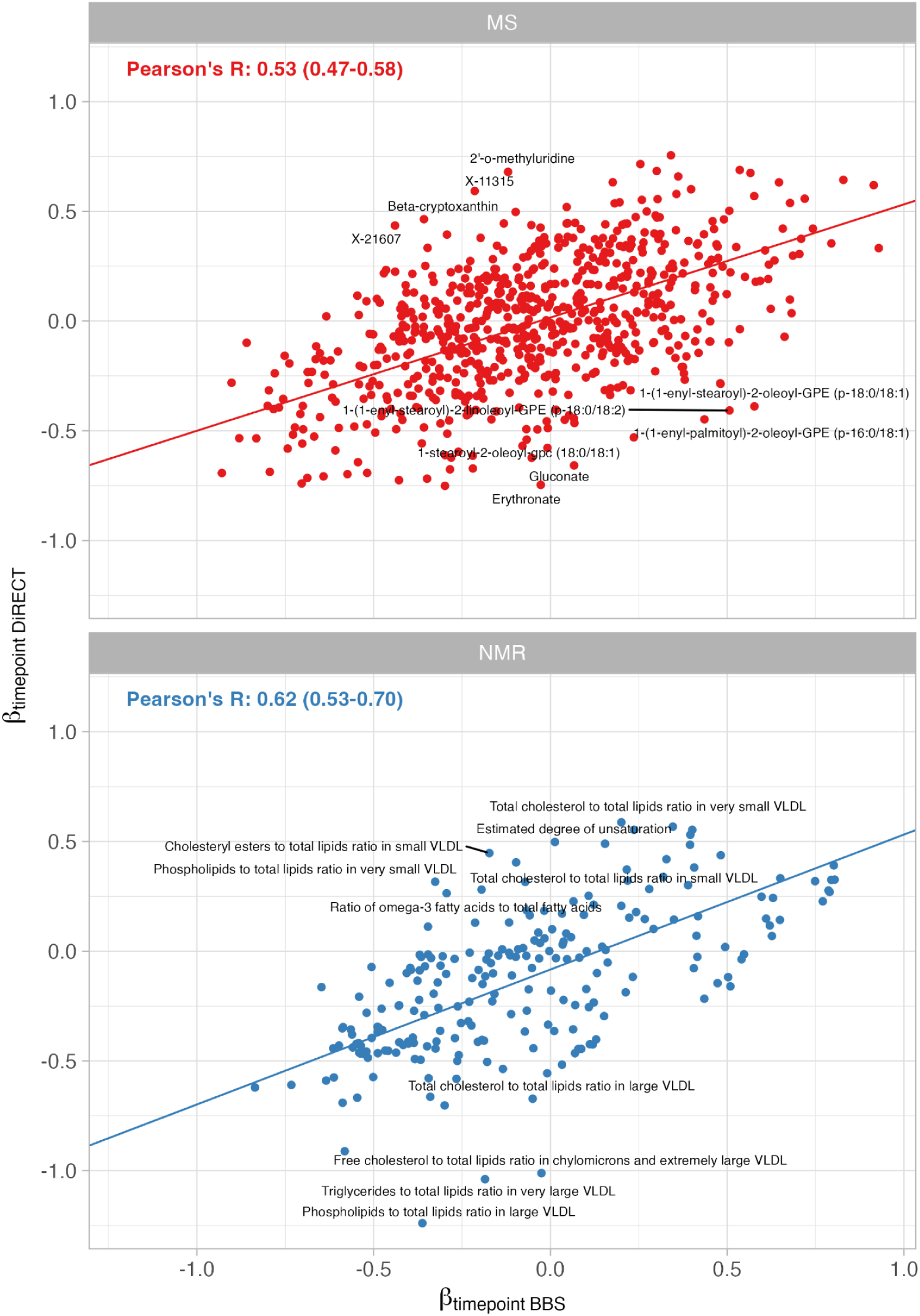
Correlation of metabolite associations between weight loss interventions. The plot presents the unadjusted (timepoint=’end’) estimates for the effect of a structured diet programme (y-axis) against bariatric surgery (x-axis) on circulating metabolite levels. The top panel presents metabolites measured on the mass spectroscopy platform (MS) and the lower panel metabolites measured on the nuclear magnetic resonance platform (NMR) platform. The x-axis represents the effect estimate and the y-axis the -log10 P value for the association. The regression lines were calculated using Deming regression to account for error in both measures. Metabolites with the greatest orthogonal distance from the regression line.

**Figure 4.**
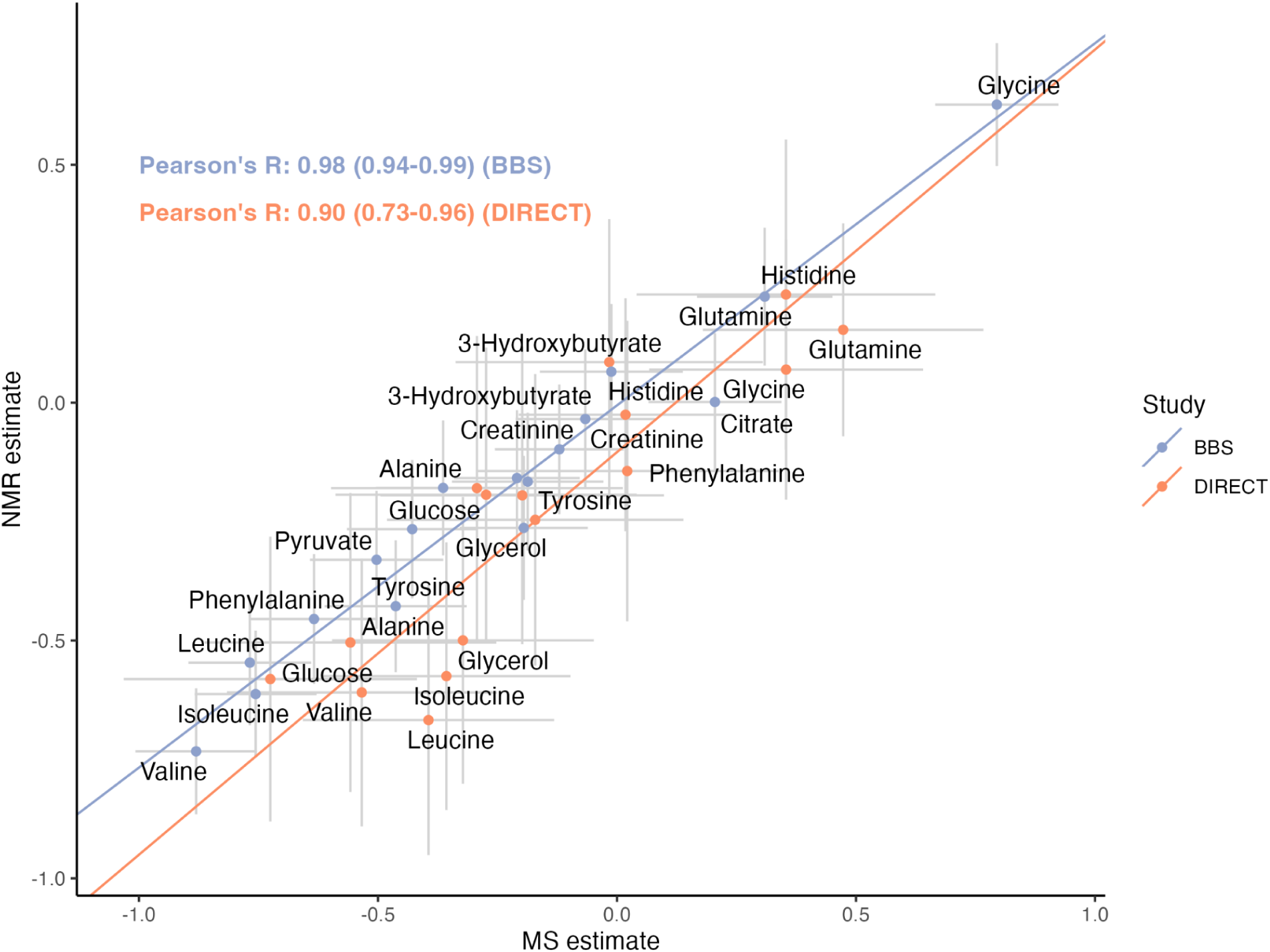
Interplatform common metabolite associations correlation. The scatter plot presents the effects for metabolites measured on both the mass spectroscopy platform (MS) and nuclear magnetic resonance platform (NMR) in each weight loss intervention study (blue, By-Band-Sleeve bariatric surgery; orange, DiRECT structured diet programme). The regression lines were calculated using Deming regression to account for error in both measures.

**Figure 5.**
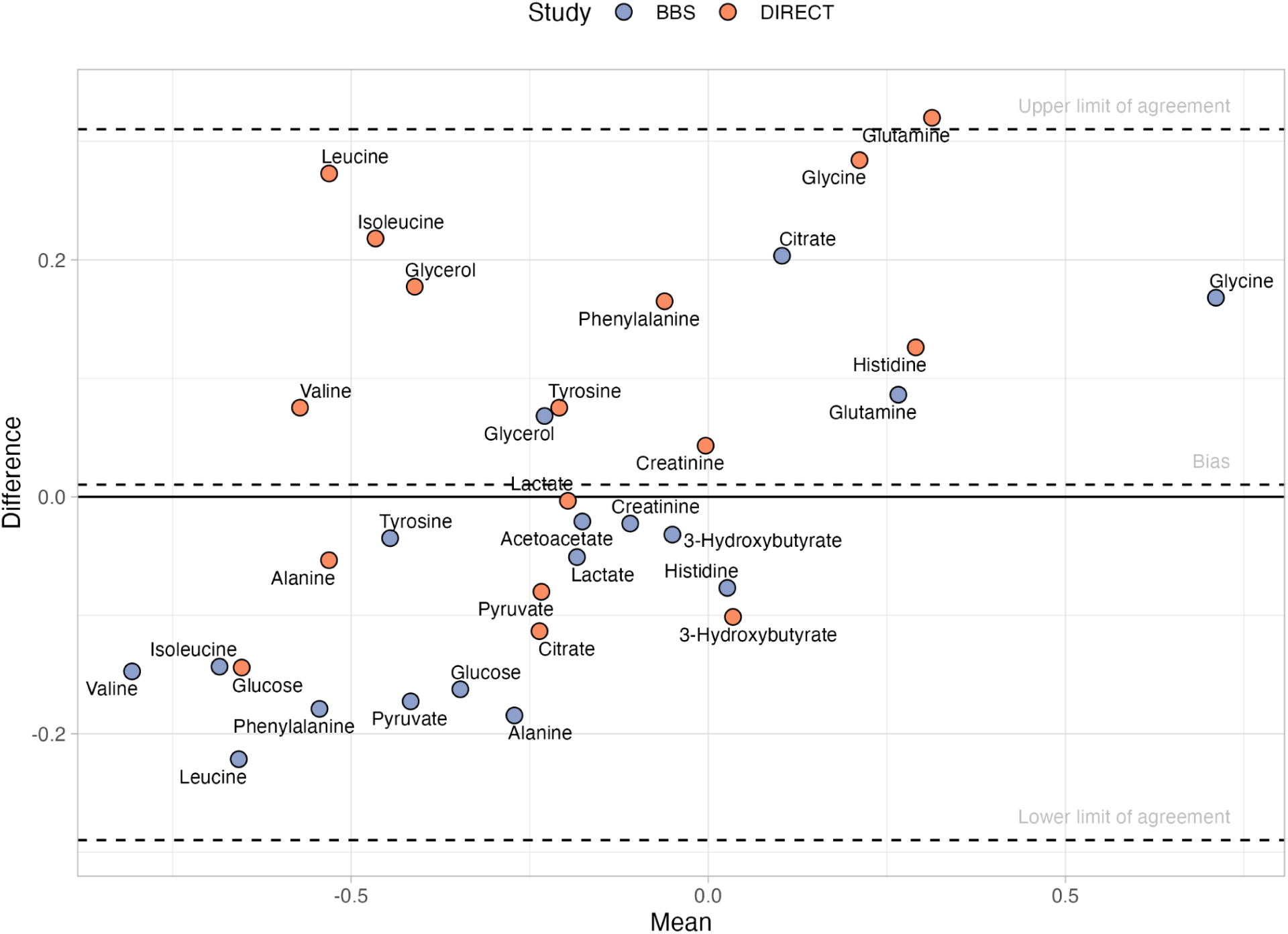
Bland–Altman plot comparing intervention effect estimates between MS and NMR metabolite measurement platforms.

**Figure 6.**
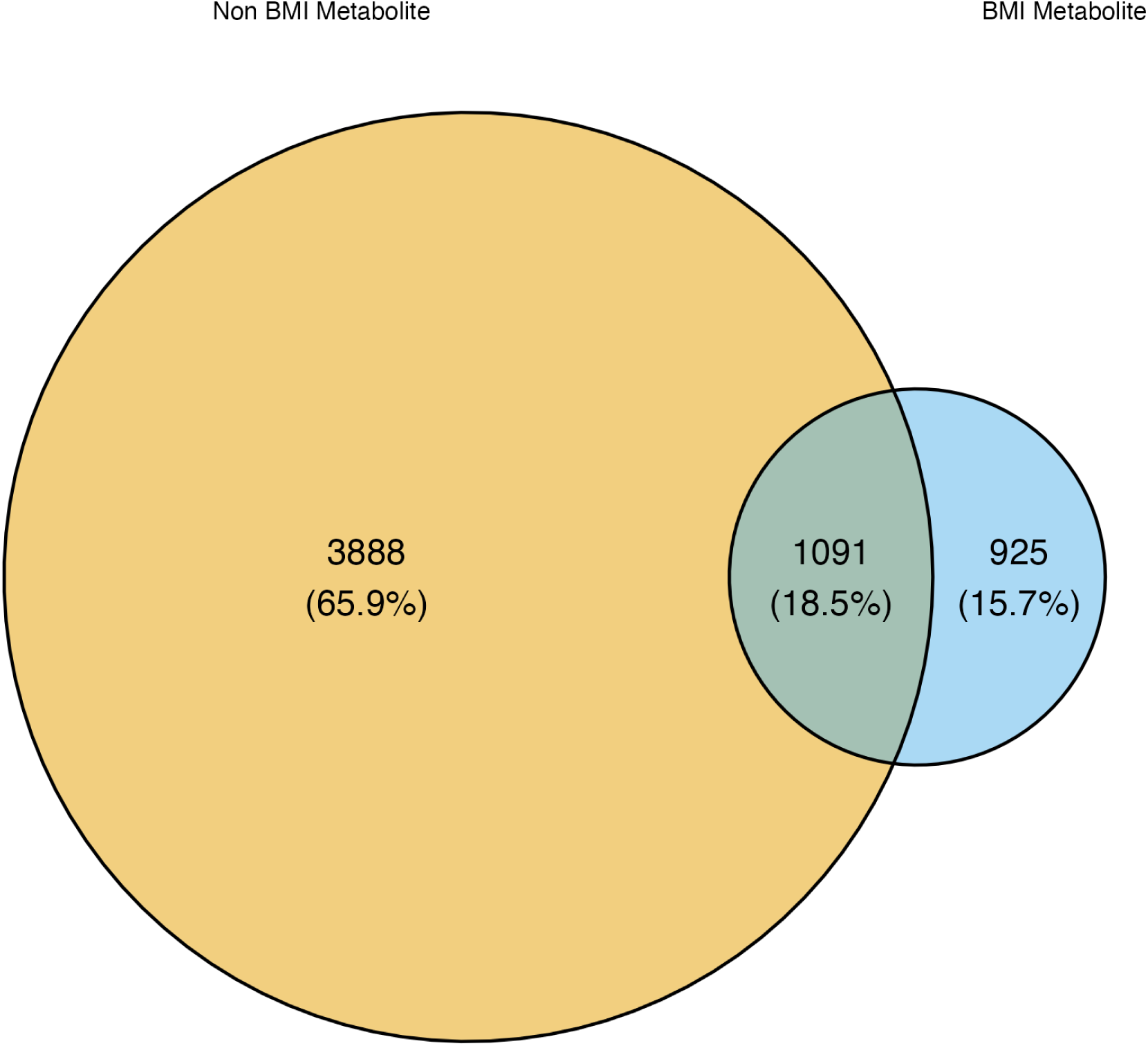
Overlap of variants between BMI-metabolite and non-BMI metabolite instruments. The Venn diagram represents all independent genome wide significant variants that were selected into one or more of the genetic instruments for metabolite levels. Blue circle, variants selected into instruments for ‘BMI-metabolites’ with consistent direction of effect and strength of association (FDR P < 0.05) across weight loss interventions and BMI-metabolite Mendelian randomisation (MR) analyses. Yellow circle, variants selected into instruments for metabolites that are not consistent across weight loss interventions and BMI-metabolite MR analyses. The overlap represents the variants that appear in both BMI and non-BMI metabolites.

**Figure 7.**
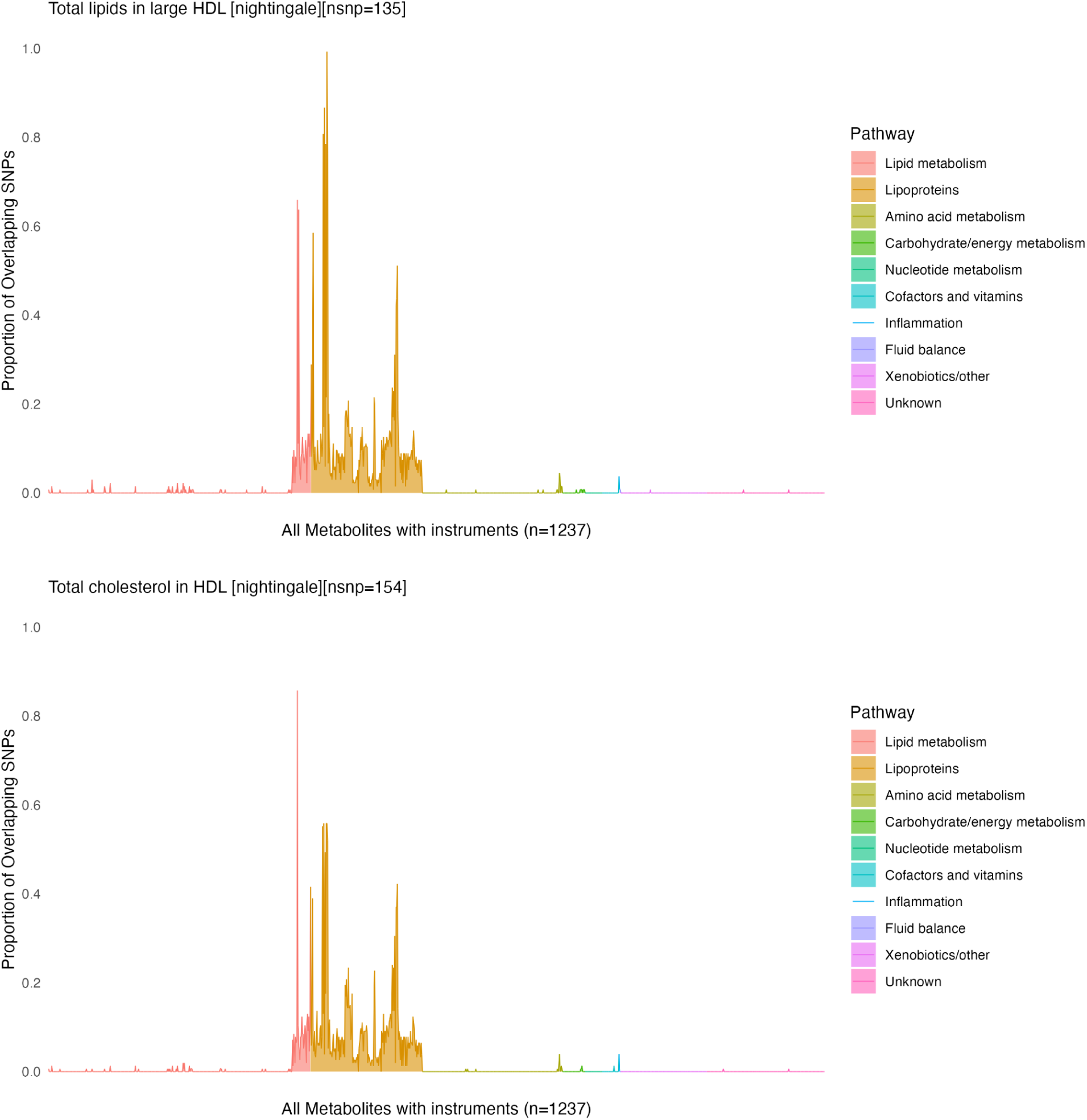
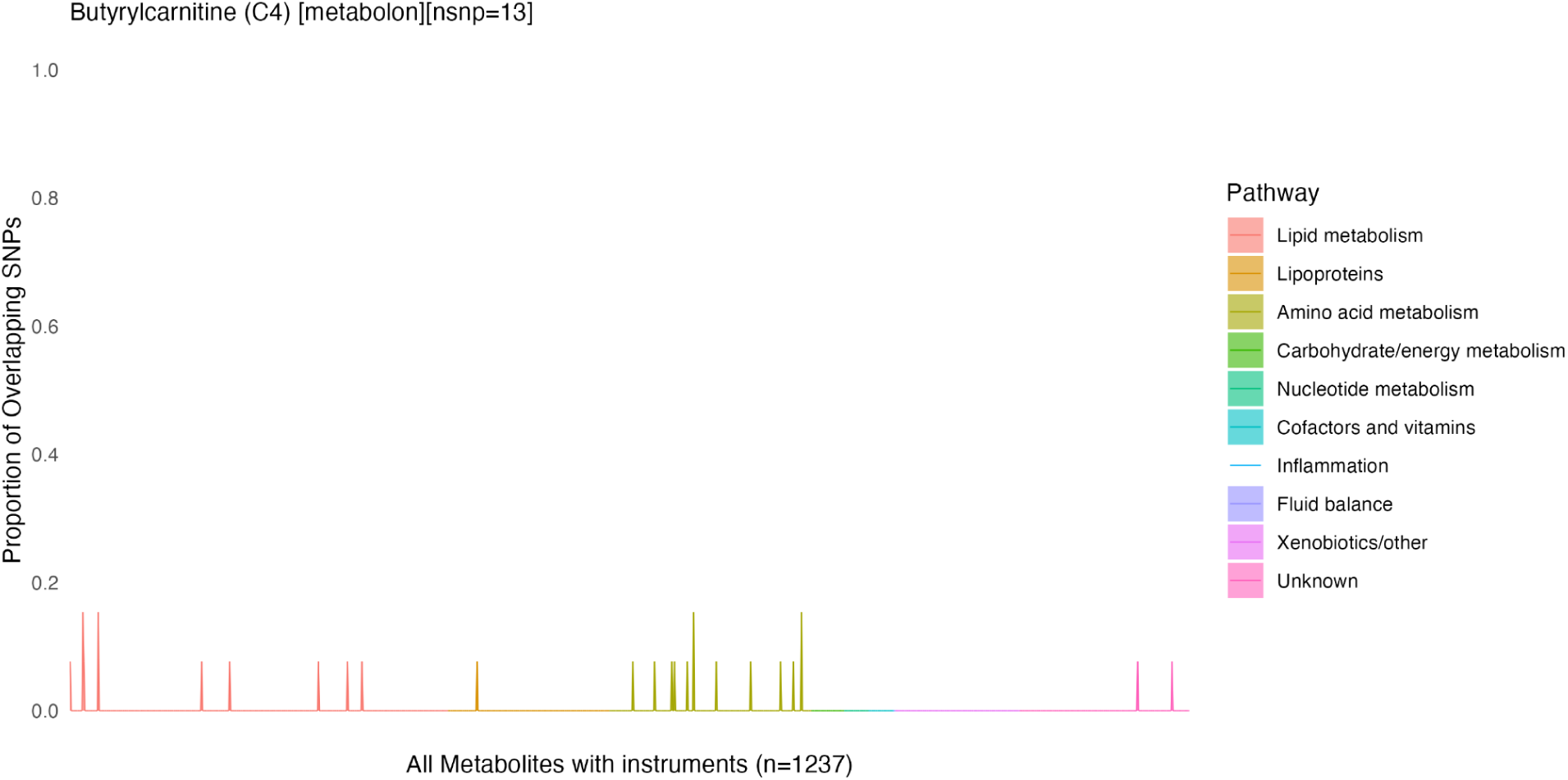
Examples of overlap between metabolite genetic instruments. The plots present the degree (%, y-axis) of overlap between variants used to instrument the metabolite and all other metabolites with a valid instrument (x-axis). The colors represent metabolite pathway groupings. The exact number of variants used in Mendelian randomization analyses may differ depending on variant presence in the outcome GWAS.

**Figure 8.**
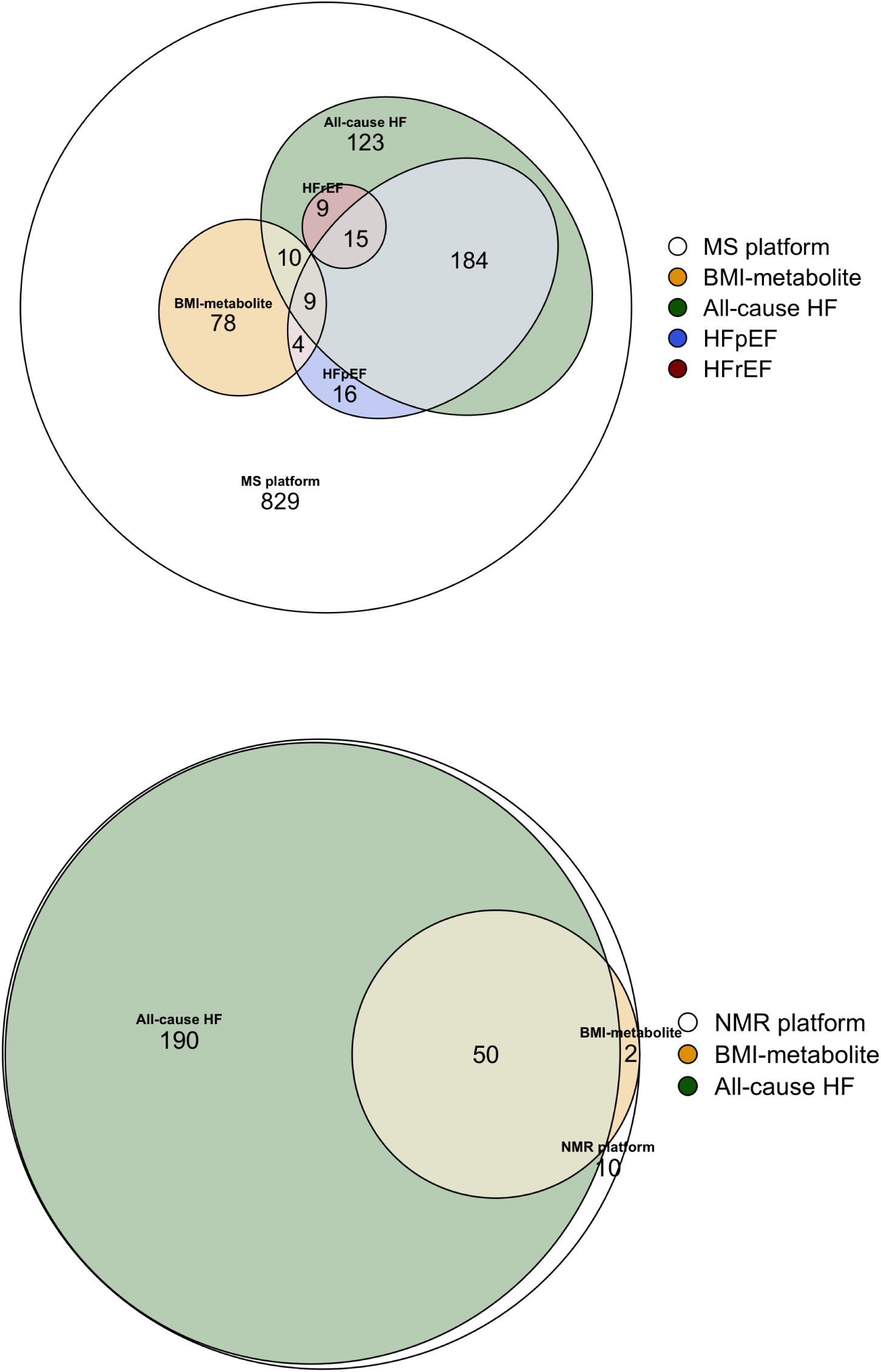
Overlap between BMI-metabolites and observational associations with heart failure.

**Figure 9.**
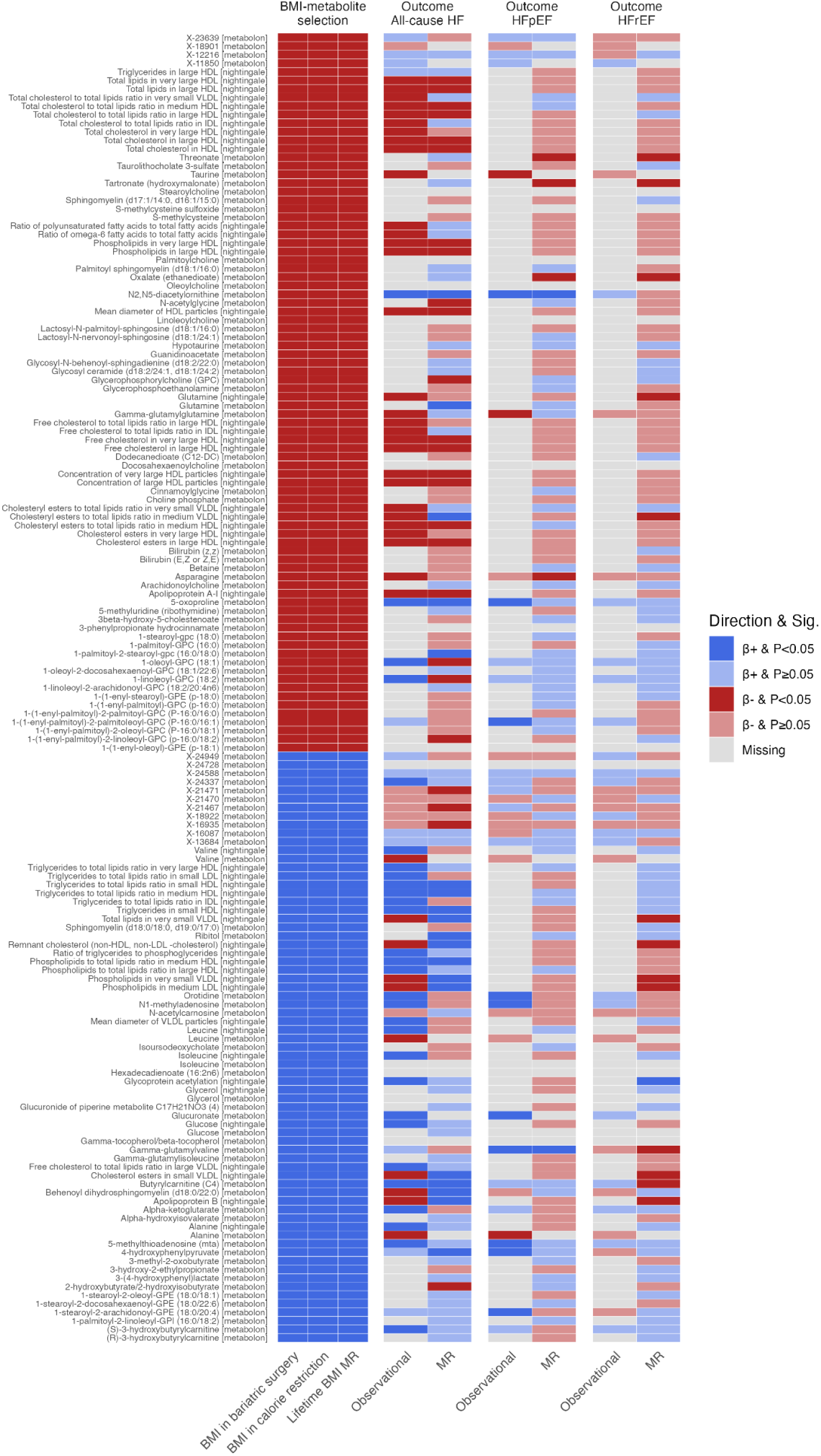
BMI-metabolites MR and observational outcome associations with heart failure.

**Figure 10.**
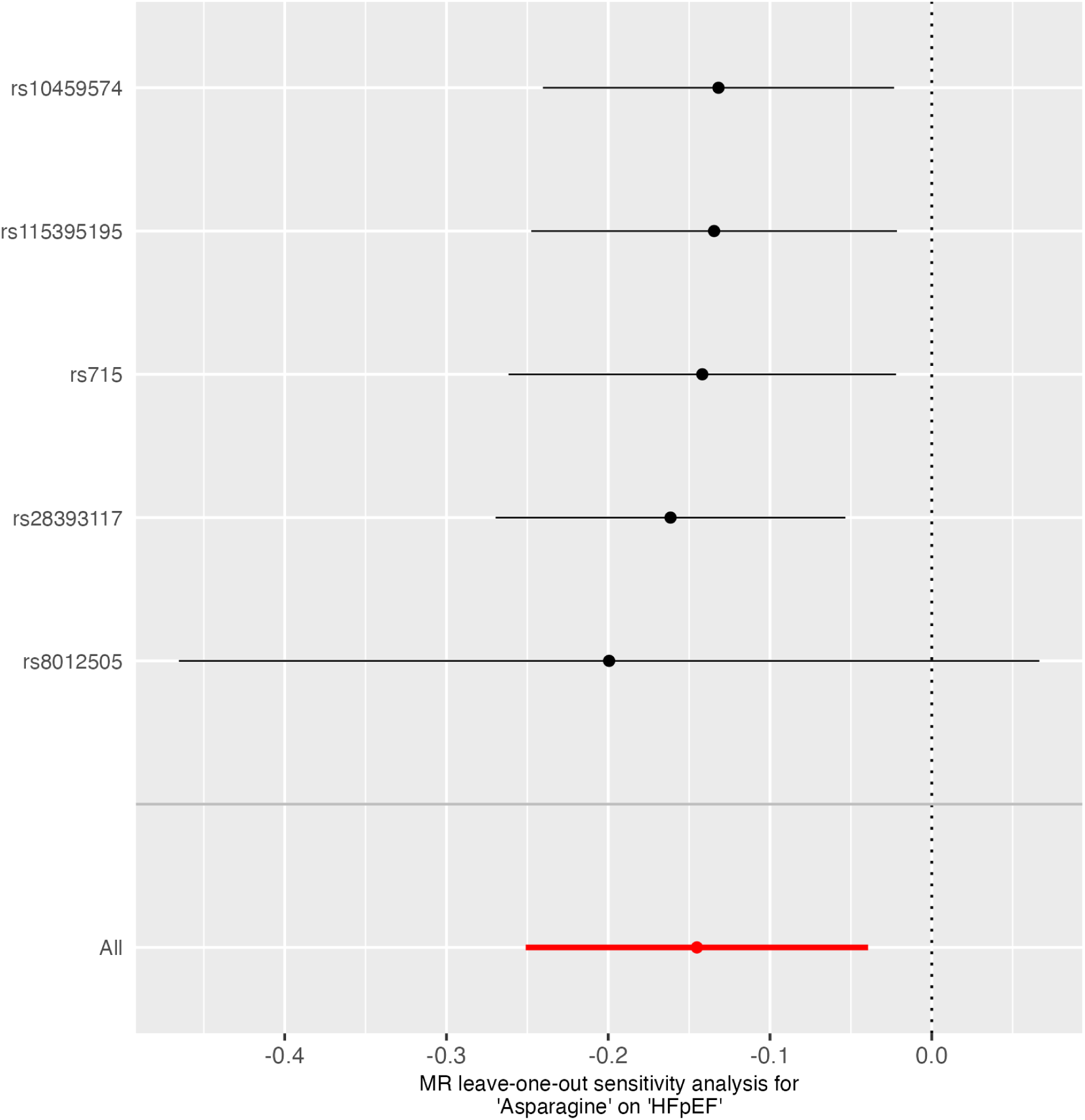
Leave-one-out analysis of the MR associations between asparagine levels and non-ischaemic heart failure with preserved ejection fraction.

